# Neighbourhood topology unveils pathological hubs in the brain networks of epilepsy-surgery patients

**DOI:** 10.1101/2024.10.02.24314429

**Authors:** Leonardo Di Gaetano, Fernando A.N. Santos, Federico Battiston, Ginestra Bianconi, Nicolò Defenu, Ida Nissen, Elisabeth C. W. van Straaten, Arjan Hillebrand, Ana P. Millán

## Abstract

Pathological hubs in the brain networks of epilepsy patients are hypothesized to drive seizure generation and propagation. In epilepsy-surgery patients, these hubs have traditionally been associated with the resection area: the region removed during the surgery with the goal of stopping the seizures, and which is typically used as a proxy for the epileptogenic zone. However, recent studies hypothesize that pathological hubs may extend to the vicinity of the resection area, potentially complicating post-surgical seizure control. Here we propose a neighbourhood-based analysis of brain organization to investigate this hypothesis. We exploit a large dataset of presurgical MEG (magnetoencephalography)-derived whole-brain networks from 91 epilepsy-surgery patients. Our neighbourhood-focus is two-fold. Firstly, we propose a partition of the brain regions into three sets, namely resected nodes, their neighbours, and the remaining network nodes. Secondly, we introduce generalized centrality metrics that describe the neighrbourhood of each node, providing a regional measure of hubness. Our analyses reveal that both the resection area and its neighborhood present large hub-status, but with significant variability across patients. For some, hubs appear in the resection area; for others, in its neighborhood. Moreover, this variability does not correlate with surgical outcome. These results highlight the potential of neighborhood-based analyses to uncover novel insights into brain connectivity in brain pathologies, and the need for individualized studies, with large-enough cohorts, that account for patient-specific variability.

## I. INTRODUCTION

The network description of complex systems such as the brain provides a remarkable tool to unveil its underlying organization and emergent dynamics. Such a description has enriched our understanding of brain organization both at the macroscopic [1–4] and microscopic [5] levels, and has found remarkable clinical applications [6, 7]. A notable example, which is the focus of this study, is the case of epilepsy surgery. This is the treatment of choice for drug-resistant epilepsy patients, and it entails the removal or disconnection of a set of brain regions –the epileptogenic zone (EZ)– with the goal of stopping seizure generation and propagation [8–10]. In practice, there is no gold standard to identify the actual EZ, instead the EZ may be approximated by the resection area (RA) in combination with surgical outcome: for patients with good outcome the EZ is included in the RA, whereas for patients with bad outcome the EZ was at least partially preserved by the surgery. Epilepsy surgery is preceded by an extensive presurgical evaluation, involving different imaging modalities such as magnetic resonance imaging (MRI) or electro- and magneto-encephalography (E/MEG). However, positive outcome rates (i.e. seizure freedom after the surgery) are not optimal, and around 30% of the patients continue to present seizures one year after the resection, although this number can go up to 50% for cohorts with complicated etiology. With the goal of improving these outcome rates, network-based studies have investigated in detail the brain network organization of epilepsy patients in order to unveil pathological effects that may predict surgical outcome [11, 12].

Within this context, a big conceptual leap has taken place, from the notion of individual epileptogenic zones, to the consideration of epileptogenic networks that arise from the interplay between different brain regions in promoting and inhibiting ictal activity [13–16]. According to this perspective, the effect of a given surgery cannot be determined alone by the characteristics of one or more regions, but needs to measured against the whole epileptogenic network [17]. Data-driven and modeling studies seem to support this hypothesis, and thus network mechanisms are recognized to participate in the generation and propagation of seizures [18–27].

Substantial evidence underscores changes in structural and functional brain networks in epilepsy [28, 29], particularly related to the EZ [30]. Whether there is an increase or decrease in connectivity of the EZ compared to healthy individuals, however, remains an open question. fMRI-based studies initially pointed towards a disconnection of the EZ [31–33], which has been supported by some invasive EEG [34] and MEG studies [35, 36]. However, recently several M/EEG studies have suggested hyperconnectivity of the EZ and neighbouring regions [14, 30, 37–43].

Pathological changes in brain connectivity in epilepsy are disproportionally associated with the network hubs [44] –highly central or important regions in the network architecture of the brain– a finding echoed in other neurophysiological disorders such as Azheimer’s disease, multiple sclerosis, or stroke [28, 45]. In the case of epilepsy, pathological hubs that facilitate seizure generation and propagation may be present. Indeed, the properties of the brain hubs, including their spatial distribution and overlap with the RA, are associated with epilepsy surgery outcome [46–51]. Notably, however, hubs can also have an inhibitory effect to prevent the ictal state [34, 50], and it should be noted that hub removal can lead to increased side-effects from the surgery. The RA and the EZ have been associated with brain hubs by several studies, both in the ictal [52–54] and interictal [40, 52, 54] states. Such studies found associations between hub removal and seizure-freedom with different MEG-based connectivity measures [40, 55], although in a recent study involving a large cohort (*n* = 91) of epilepsy-surgery patients we could not confirm these findings [41]. In a recent MEG study with a smaller cohort of 31 epilepsy patients, [43] were able to classify epilepsy surgery patients according to surgical outcome (79% accuracy and 65% specificity) by comparing the degree centrality (a measure of *hubness* given by the number of neighbours of a node) of the RA to the remaining network node, in the pre-surgical brain-network.

Overall, although hub removal has been associated with a favorable outcome of epilepsy surgery, this does not seem to be a necessary condition for a good outcome. Indeed, brain hubs do not always overlap with the RA, even for seizure-free patients [20, 21, 23, 41]. These findings motivated the hypothesis that the seizure onset zone (SOZ), (the region where seizures start), need not coincide with the pathological hubs but may be strongly connected to them, in which case removal of either the SOZ, the pathological hub, or even the connection between them may be enough to prevent seizure propagation and achieve a good outcome [20, 41]. Thus, regional brain organization around the SOZ –as opposed to only its centrality– becomes a promising target to understanding the effect of a given resection.

In order to study the role of regional brain organization in epilepsy surgery, we propose here a neighbourhood-based description of brain connectivity and of the effect of a resective surgery in epilepsy. Firstly, we consider explicitly the role of the connectivity of the RA-neighbours by implementing a partition of the brain regions, namely into RA-nodes, its neighbourhood, and the remaining network. By doing so, we are able to specifically address the question of the emergence of pathological hubs in the vecinity of the RA, and their relation to surgical outcome. Secondly, we introduce a novel analysis framework to quantify regional brain organization based on the notion of extended neighbourhoods, following a previous theoretical study that generalizes the notion of clustering coefficient [56]. The extended neighbourhood of a node describes its area of influence, providing a mesoscopic description of brain organization that can inform us of e.g. the existence of regions with strong recurrent connectivity [56]. By characterizing the extended neighbourhood of each brain region through topological data analysis [57], we propose the generalization of local node-based centrality metrics to regional descriptors that encode regional organization. As we go on to show, the neighbourhood-based description unveils the distinctive properties of the neighbours of the RA. In this multi-frequency study (including 6 specific frequency bands as well as the broadband) we found that the RA and its neighbours shared a highly central status that was significantly larger than that of the remaining brain regions. The relative centrality of the RA and its neighbours varied within the population (and between frequency bands and network metrics), and whether the RA or its neighbours were more central was not an indicator of surgical outcome (Area Under the Curve *AUC* = 0.46 for the classification of patient outcome for the broadband network). In contrast, we achieved a fair classification of the patient groups (*AUC* = 0.62, 0.64 respectively) when considering either the hub-status of the RA or of its neighbourhood in relation to the remaining brain regions. These findings support the notion of the emergence of pathological hubs in the brain of epilepsy patients that may not coincide with the seizure onset zone but appear in its neighbourhood. Overall, our findings highlight the need to consider regional brain connectivity in epilepsy surgery studies, for instance with the notion of node neighbourhoods as proposed here.

## II. RESULTS

### A. Extended Neighbourhoods

In order to characterize regional brain organization we have considered the notion of the extended neighbourhood *ε*𝒩 of a node [56]. Extended neighbourhoods, also called ego-centered networks, define the area of influence of a node. Mathematically, the extended neighbourhood of node 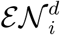, is defined as the subgraph conformed by nodes at distance *δ*, 0 *< δ* ≤ *d*, of node *i* (which, crucially, excludes node *i*, see Methods for a detailed definition), as depicted in figure 1. By changing the radius *d* of the extended neighbourhood we can access different scales of network organization, going from the local to the global perspective. In order to quantify the structure of 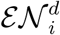, and thus regional network organization, we have considered five topological measures: the size or number of nodes 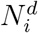, the number of edges 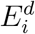, and the first three Betti numbers quantifying the number of connected components 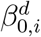, the number of loops 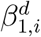 (not accounting for triads which are always considered to be filled, see Methods), and the number of cavities or 2-dimensional loops, 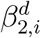. These metrics quantify the topology of the extended neighbourhood of each node. The number of nodes and edges indicate the regional connectivity of the node, and can be interpreted as centrality metrics. Similarly, a high value of the first Betti number indicates that node *i* acts as a broker between different otherwise disconnected components of its neighbourhood [56]. In figure 2 we provide an illustration of these different metrics. A more detailed description of each metric can be found in the Methods and in the Supplementary Material.

**FIG. 1:**
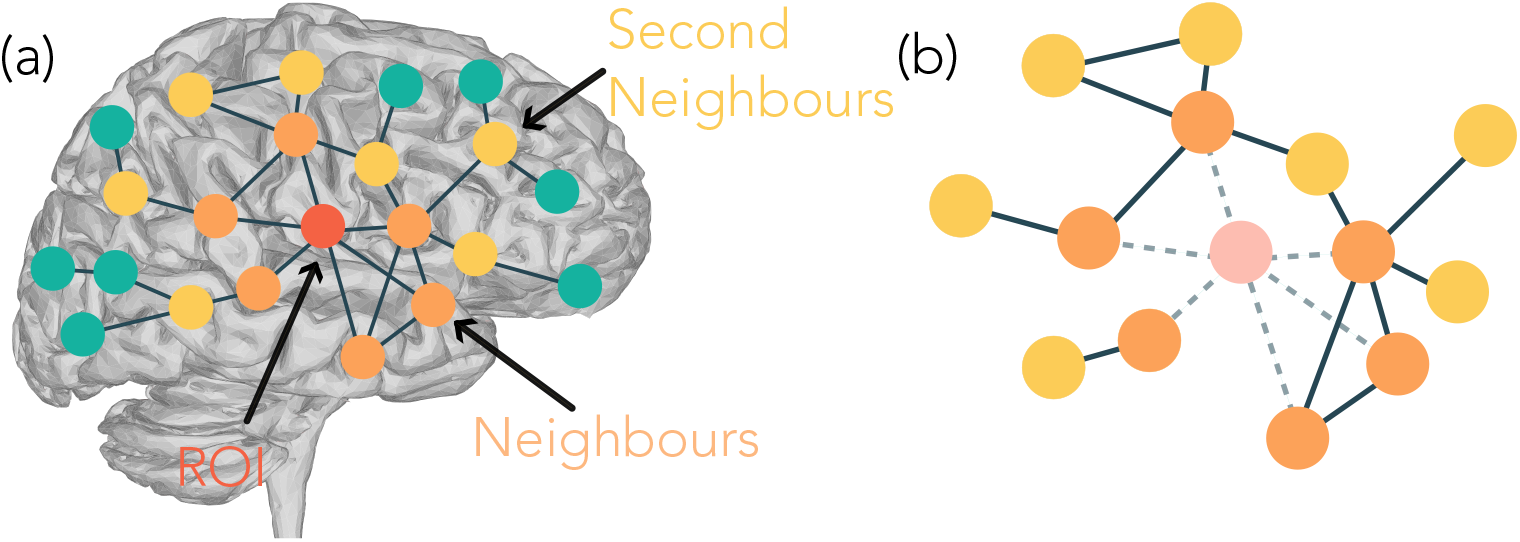
Schematic description of extended neighbourhoods. (A) Illustrative representation of the extraction of a node’s neighbourhood for *d* = 2. The nodes are color-coded to show the central node (red), its first (orange) and second (yellow) neighbours and the remaining network nodes (green). (B) Extended neighbourhood of the node. The central node is not included in its neighbourhood, therefore it is shown here with low opacity (light pink node) and its edges are removed (dashed lines). The topological organization of the neighbourhood can be observed. In this case, e.g. two different connected components emerge, as well as two closed triangles.

**FIG. 2:**
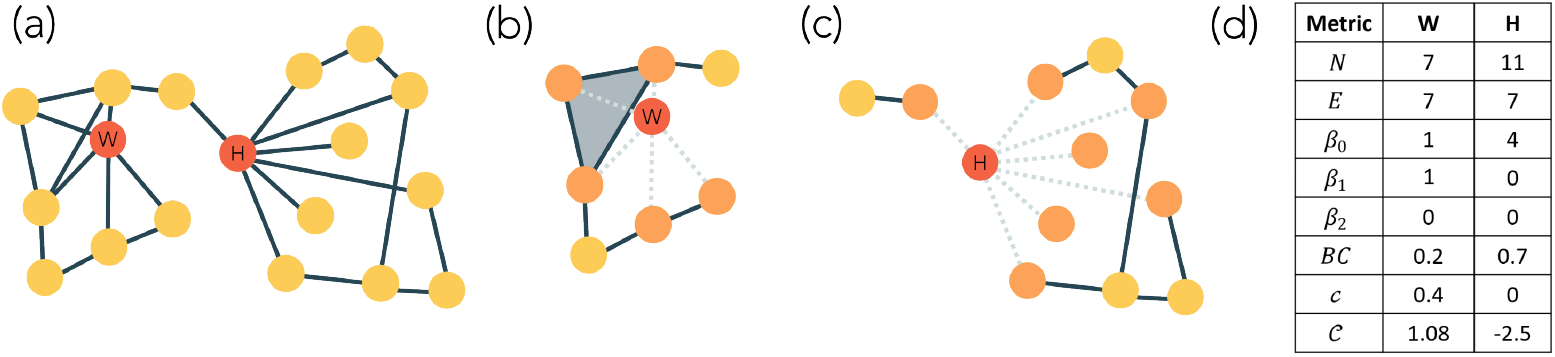
Illustration of the properties of simplicial complexes and extended neighbourhoods. (a) Schematic network were we highlight two nodes: a regional hub (node *H*) with high degree (7 neighbours) and high *BC* since it brokers two communities, and a local hub (node *W*) with high degree (5 neighbours) but low *BC*. Panels (b) and (c) illustrate extended neighbourhoods of *W* and *H*, respectively. The different topology of *ε*𝒩 _*W*_ and *ε*𝒩 _*H*_ is encoded by the regional and local metrics, as shown in panel (d). Whenever there is a closed clique in the original network, simplices are built in the extended neighbourhood. For instance, in panel (b), the grey triangle represents a 2-dimensional simplex built according to this rule.

As a benchmark, we have considered also three node-based metrics, namely the betweenness centrality BC_i_, the local clustering coefficient c_i_, and the local curvature C_i_. The betweenness centrality is a standard measure to quantify node-centrality and define hubbness [20, 28, 40]. It quantifies the extent to which a node lies on the shortest paths between other nodes, thus capturing its role in controlling information flow in the network [58]. Node curvature measures how paths in the simplicial complex diverge or converge around a node, capturing the local geometric properties of the space. Specifically, in a simplicial complex, curvature reflects how higher-dimensional simplices (such as triangles or tetrahedra) connect around a node, influencing the shape and flow of the network structure. It is associated with network robustness, and also identifies brain hubs, with large negative values being indicative of hub status [59]. The clustering coefficient captures the connectedness of a node’s neighbours, and has previously been associated with epilepsy surgery outcomes [19].

### B. Topological characterization of the epileptogenic zone

Following the hypothesis that the EZ is either a hub or connected to a hub, we hypothesized that RA nodes and their neighbourhoods will be more central than other nodes in the network. In order to test this hypothesis, we considered an existing database comprising 91 patients who underwent epilepsy surgery at Amsterdam UMC, location VUmc. This database had been studied with a combination of network metrics and machine learning previously [41]. The brain organization for each patient was encoded in a functional brain network comprised of 90 regions of interest (ROIs) (according to the AAL atlas [60]), derived from resting-state MEG, and thresholded to keep only the strongest links (see Methods for details). MEG networks were derived in different frequency bands, which account for different aspects of brain function. For simplicity we have considered here first the broadband (0.5 − 48.0Hz), but refer back to a multi-frequency analysis in later sections. The resection area of each patient was derived from post-operative MRI and was encoded in terms of AAL nodes.

Each node in the network was described by means of the 8 metrics defined in the previous section, with high values of these metrics associated with higher generalized centrality, except for the curvature where the direction is the opposite as discussed above. Initially, two sets of nodes were defined for each patient and network: resected nodes ℛ𝒜 and non-resected nodes 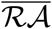. We analysed whether these nodes differed at the individual level in any of the 8 metrics considered (bootstrapping and a Bonferroni correction were used to establish statistical significance, see details in the Methods). Details of this analysis for an exemplary case are shown in Supplementary Figure 2. We found that, at the individual level, ℛ𝒜 nodes were significantly more central according to all neighbourhood metrics, except *β*_0_, for 15% to 30% of patients (respectively for 23, 27, 20 and 14 cases for *N, E, β*_1_ and *β*_2_). Traditional node-based metrics were less efficient at detecting differences between the node groups: according to these metrics ℛ𝒜 nodes were significantly more central than 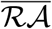 nodes only for a handful of patients (respectively 6, 3 and 6 for *c, C* and *BC*). We note that, for a few patients, the opposite result was found and ℛ𝒜 nodes were significantly less central than 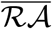 nodes, both with the node- and neighbourhood-based metrics (respectively 5, 5, 7 and 3 cases for *N, E, β*_1_ and *β*_2_; and 3 and 1 cases for *c* and *C*; whereas no case was found for *BC*). These results are summarized in figure 3 (a), whereas numerical results can be found in Suppl. Tables 2 and 3.

**FIG. 3:**
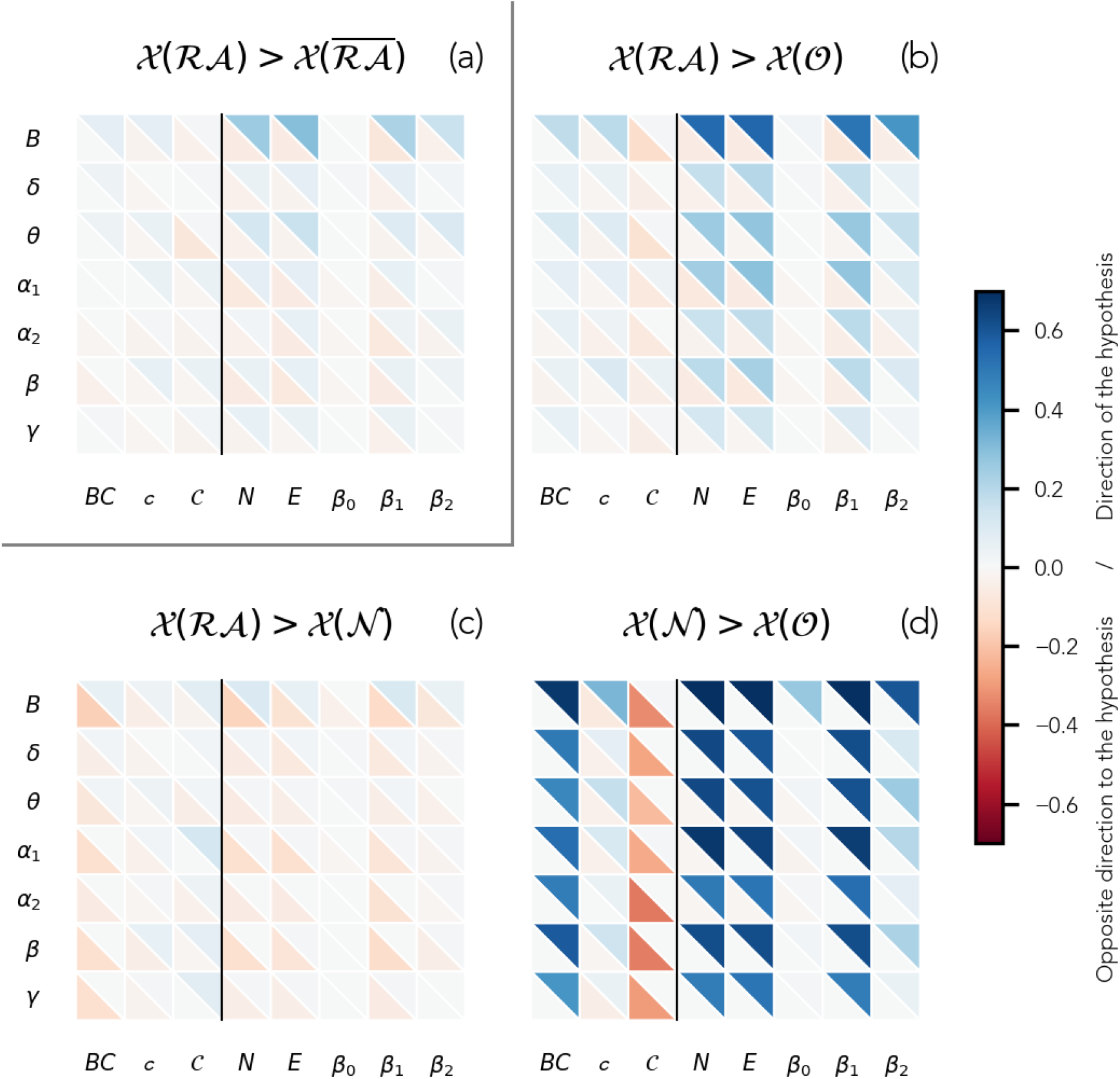
Patient-specific comparison of different node groups for the two-group (a) and the three-group (b, c, d) set-ups. For each panel, the hypothesis of the relation in centrality between the two metrics is shown in the panel title. The fraction of patients for whom there was a significant difference in the direction (opposite direction) of the hypothesis is shown by the blue (red) triangles in the upper-right (bottom-left) corner of each cell, respectively for each frequency band (rows) and metric (columns), color-coded as indicated by the colorbar. The corresponding numerical values are shown in Supp. Tables 2 and 3. The vertical black line separates node-based (left) from neighbourhood-based (right) metrics. 𝒳 (*S*) stands for the generalized centrality metric 𝒳 measured on the nodes in set *S*.

Our results agree with previous studies according to which the 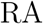 is not always a network hub, but it is often strongly connected to a pathological hub [20, 28, 41]. Consequently, the 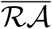 node set may include both nodes that are less and more central than ℛ𝒜 nodes. In order to take into account this effect, we split the 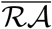 set into two: nodes that were neighbours of the ℛ𝒜 (*neighbours*, 𝒩 set) and nodes that were not (*other*, 𝒪 set). According to our initial hypothesis, within this division of the node sets we expected that both ℛ𝒜 and 𝒩 nodes were more central than 𝒪 nodes, and that ℛ𝒜 and 𝒩 nodes were similarly highly central. As expected, we found that the node sets ℛ𝒜 and 𝒩 were in most cases significantly more central that the 𝒪 set (see figure 3 panels b and d). As before, neighbourhood-based metrics were able to capture this difference more consistently across patients than node-based metrics. Regarding the relative hub-status of the ℛ𝒜 and its neighbourhood, we only found significant differences between ℛ𝒜 and 𝒩 nodes for a small fraction of the patients (figure 3 panel c). These went in both directions, with a tendency towards a higher centrality of 𝒩 nodes at the group level, as we discuss below. For instance, for the metric that picked up the most differences in the broadband, *β*_1_, ℛ𝒜 nodes were more central than 𝒩 nodes for 10 cases, but the opposite was true for 12 cases. These findings indicate heterogeneity in the patient population regarding the relative hub-status of the resection area and its neighbours. For most cases, these two sets could not be distinguished based on centrality metrics (either node-or neighbourhood-based), indicating a similar highly-central status (note that the remaining nodes were found to be less central).

#### 1. Group level analyses

In order to gain a population-level perspective of the relative hub-status of the RA, we repeated the previous analyses at the group level. To do so, we measured the average centrality of the nodes in each node-set, for each patient and frequency band. We found that, when all patients were pooled together, the differences between node-sets became more subtle, likely due to patient-specific variability, as shown in figure 4. Overall, we found in the two node-group analysis that the ℛ𝒜 and 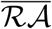 node-sets could not be significantly distinguished at the group level, for most metrics and frequency bands, with the most notable exception of the broadband. The three node-group analysis recovered for the most part the findings of the individual-level analyses, i.e. 𝒪 nodes were the least central, and 𝒩 were somewhat more central than ℛ𝒜. At the group level the betweeness centrality became the most robust metric across frequency bands, and the broadband network was the network for which differences between node-groups were more prevalent across metrics. Notably, three of the metrics, the local clustering *c, β*_0_ and *β*_2_, performed poorly for the remaining frequency bands.

**FIG. 4:**
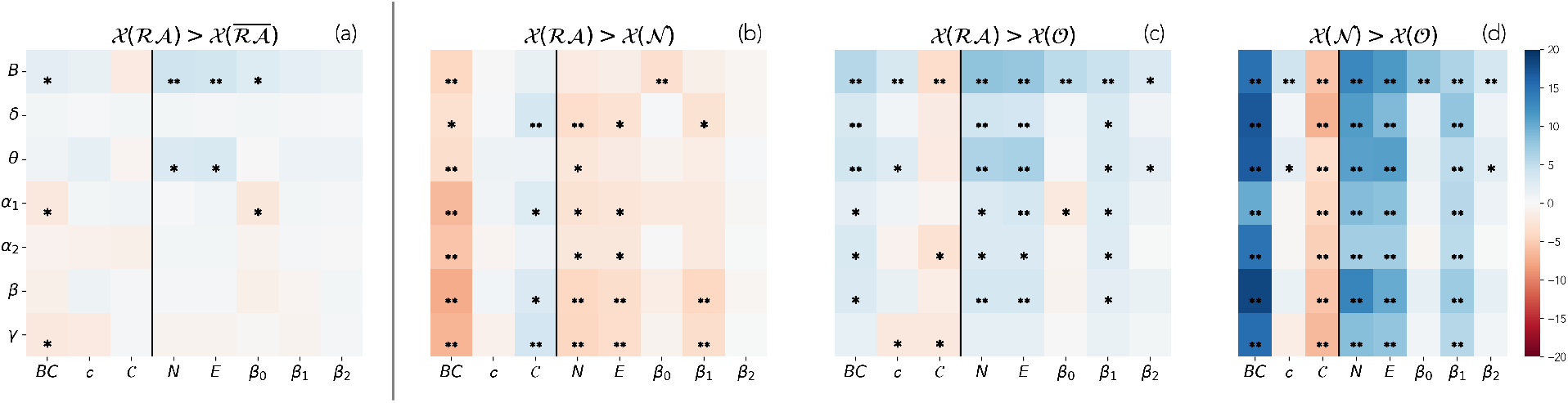
Group-level comparison between nodes sets, for each considered frequency-band (y-axis) and network metric (x-axis). From left to right, the panels indicate the difference between the node sets: ℛ𝒜 vs 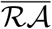, ii) ℛ𝒜 vs 𝒩, iii) ℛ𝒜 vs 𝒪, iv) 𝒩 vs 𝒪. 𝒳 (*S*) stands for the generalized centrality metric 𝒳 measured on the nodes in set *S*. The color code indicates the *z*-score of the difference between the average values of each node set, computed by bootstrapping the data (sampling size of 10^4^). Single asterisks indicate significant differences (*p <* 0.05) that did not survive the Bonferroni correction (*n* = 56, see Methods), and double asterisks the ones that did (*p <* 8.9^−4^). The corresponding numerical values are shown in Supp. Tables 4 and 5.

### c. Topological signatures of the RA and surgical outcome

In order to investigate whether the hub-status of the ℛ𝒜 was associated with surgical outcome in this dataset, we assigned each patient a *distinguishability* score 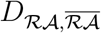 to quantify the distinguishability between the ℛ𝒜 and 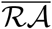 node-sets [43]. For each patient, 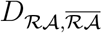 measures the number of tests (over the 8 network metrics considered) for which the hypothesis of the hub-status of the RA is significantly fulfilled (see Methods for details on data and how the statistical analyses have been performed.). Following our previous findings that a three-node-group division is more informative at the node level, we also assigned distinguishability scores to the pairwise comparisons between the three node-sets ℛ𝒜, 𝒩 and 𝒪, namely *D*_ℛ𝒜,𝒩_, *D*_ℛ𝒜,𝒪_ and *D*_𝒩,𝒪_. Next, to summarize the results of the three-node-group analysis into one score, we defined a combined score of the three-node-group analysis *D*_comb_ by summing over the corresponding three pairwise comparisons (see Methods for details). The derivation of these metrics is illustrated in Supplementary Figure 3. We used each distinguishability score to classify the patients between the *seizure-free* (SF) and *non-seizure-free* (NSF) groups, as shown in figure 5 (a statistical comparison between the two groups was also performed, see Supplementary Figure 4, but no statistical differences between the two groups survived after Bonferroni correction). We found that patient classification was fair at best for any of the frequency bands or node-group montages (panel c). The best results were found for the broadband when considering the combined information of the three-node-group analyses, which resulted in an area under the curve *AUC* = 0.68.

**FIG. 5:**
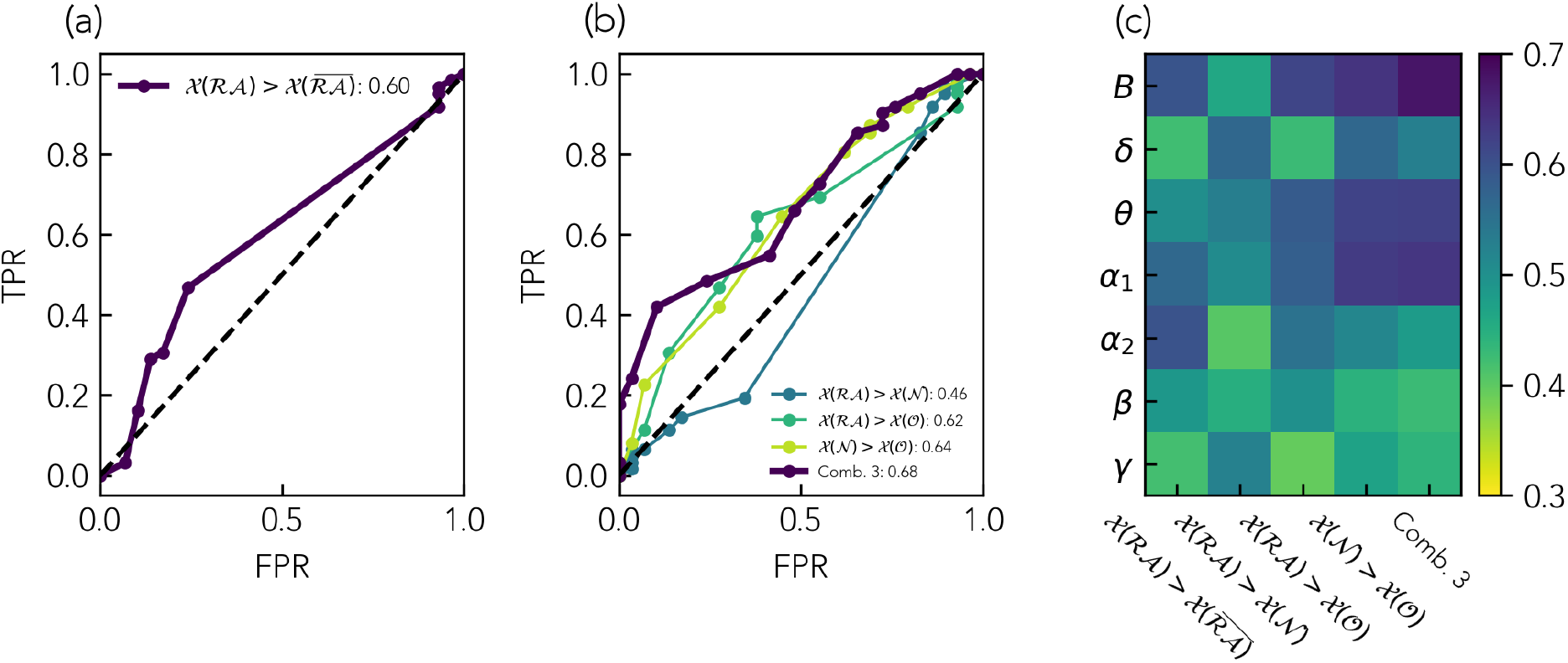
Classification of SF and NSF patients based on the two-node-group (a) and three-node-group (b) distinguishability scores. 𝒳(*S*) stands for the generalized centrality metric 𝒳 measured on the nodes in set *S*. Panels (a) and (b) show the ROC curves corresponding to the broadband, the remaining bands are shown in Supplementary Figure 5. The resulting AUC is indicated by the figure legends. Panel (c) shows the resulting AUC for all frequency bands, for this same analysis. In this representation, the SF group is assigned to be the positive class. The color-scale is centered around *AUC* = 0.5, which indicates a lack of association. Blue-colors stand for an association in the direction of the hypothesis (*AUC >* 0.5, i.e. the SF group presents a higher distinguishability score) whereas red-colors stand for the opposite (*AUC <* 0.5, the NSF group presents a higher distinguishability score).

Finally, to better contextualize and validate our findings, we considered an alternative definition of the node distinguishability, *D*^*′*^, as introduced by [43]. In this case *D*^*′*^ is simply the area under the curve resulting from the classification of ℛ𝒜 and 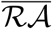 nodes (see Supplementary Figure 6). In their original study, [43] found that they could classify the patients according surgical outcome with an AUC of 0.76 using *D*^*′*^ based on the degree-centrality as metric. For our dataset, however, we found an AUC of only 0.65 when using *D*^*′*^ based on the degree-centrality (see Supp. Table 6). When applying *D*^*′*^ to the 8 metrics considered in the main part of this study, we found *AUC* values ranging from 0.68 (for the neighbourhood metric *N* in the *α*_1_-band) and 0.36 (neighbourhood metric *β*_1_, *α*-band) for the two-node-sets analysis, with similar results also for the three-node-set partition (see Supp. Figure 7).

## III. DISCUSSION

In our study involving 91 patients who underwent epilepsy surgery, we investigated the hub-status of the resection area and its region of influence to shed new light onto the presence of pathological hubs in the brains of epilepsy-surgery patients and their role in the outcome of epilepsy surgery. We proposed a novel methodology based on node-neighbourhoods and topological data analysis to quantify node centrality at a mesoscopic level. As a validation of our novel approach, we compared our findings against established node-based metrics such as the betweenness centrality and clustering coefficient. Moreover, by leveraging the same database previously analyzed by [41] with traditional methodologies, we enabled a direct comparison between the two studies.

In our study, we found that (a) the neighbours of the resection area play an important role in brain-network organization in epilepsy and are significantly different from the remaining nodes in the networks (thus a three-group partition of the brain regions, where RA neighbours are separated from the remaining brain network, is more representative than a two-group partition); (b) the RA and its neighbours are more central than the remaining brain regions, which holds true at the group level and also individually for most patients; (c) the RA and its neighbours are similarly highly-central, with only some differences at the individual level (for 10 to 20% of patients) that go in both directions, whereas at the group level the neighbours are weakly but significantly more central; and (d) the difference in hub-status between either the RA or its neighbours and the remaining network nodes, but not between them, is weakly associated with surgical outcome (*AUC* = 0.62, 0.64 and 0.46, respectively). A main consequence of our findings is that a three-node-group partition of the brain regions as we have introduced here, such that the RA-neighbouring regions are separated from the remaining brain regions and considered specifically, is more representative than a two-node-group partition, in particular yielding better node-classification results. These findings support the hypothesis of the emergence of pathological hubs in refractory epilepsy that do not necessarily overlap with the RA, a finding that was valid for patients with good and bad outcome. These results further highlight the need for individualized studies that take into account patient-specific brain connectivity.

### A. Hub status of the RA

In this study we considered the emergence of pathological hubs in epilepsy and their overlap with the resection area [28, 40]. The RA has been associated with brain hubs both in functional and structural studies ([14, 21, 38–43, 52–54, 61, 62]; see also [28, 63] for recent reviews), and their overlap has been related to surgical outcome, with several MEG studies finding that hub removal was associated with good postsurgical outcomes [40, 42, 43, 62]. In particular, [40] found that the brain network hubs (defined via the betweenness centrality on a minimum-spanning-tree, MST, description) were localized within the resection cavity in 8 out of 14 SF patients and none (out of 8) NSF patients (73% accuracy). Similarly, [62] found that removal of the most central hubs (defined via the eigenvector centrality on weighted PLI networks) had predictive value in a study with 31 patients (17 SF). Considering a simple correlation metric as the basis for connectivity, [43] found, in a study with 31 patients (12 SF), that SF patients had significantly more hubs surgically removed. Finally, [42] also found higher functional connectivity (defined via both the amplitude-envelope coupling and phase-locking-value on the MST description) inside than outside the RA for SF patients, and few differences between the two for NSF patients in a study with 37 (22 SF) patients involving both children and young adults with refractory epilepsy. The functional connectivity measures predicted weakly the EZ location and surgical outcome (sensitivity and specificity above 0.55 with leave-one-out cross-validation).

However, the relationship between hub-removal and surgical outcome could not be validated in our previous study [41] (94 patients, 64 SF) which used the same patient cohort as we have considered here. Nissen and colleagues defined the hub-status on the basis of the MST betweenness centrality, and only a weak association with the RA was found (60.34% accuracy with a random forest classifier) and none with surgical outcome (49.03% accuracy). In line with the suggestion that the relationship between the RA and the brain hubs is not straightforward, several studies have pointed towards the functional isolation of the EZ, both in invasive EEG [34] and MEG [35]. In particular, [35, 36] found that SF patients presented a more isolated resection area (relative to the contralateral hemisphere) than NSF patients in a study with 12 patients (7 SF) based on amplitude-envelope-correlation networks. [34] found that the seizure onset zone (SOZ) and the early propagation zone presented increased inwards and decreased outwards functional connectivity in an invasive EEG study involving 81 drug-resistant epilepsy patients undergoing presurgical evaluation. Interestingly, they found that the largest difference between SF and NSF patients appeared in the propagation zone: the connectivity profile of the propagation zone was intermediate to that of the SOZ and the remaining networks for SF patients, whereas for NSF patients it consistently and closely resembled that of the remaining network. It is worth noting that this result may just reflect a difference in invasive EEG sampling between SF and NSF patients, such that e.g., the true propagation zone of NSF patients may have been undersampled [34].

The existence of pathological hubs can reconcile these findings: a pathological hub that may or may not coincide with the SOZ may be present facilitating seizure propagation. Then, removal of either the SOZ, the pathological hub, or even the connection between them, could lead to seizure freedom [23, 40]. In a previous modeling study, for instance, we found that the link-based resections that led to the best postsurgical outcome in the model were those linking the RA to the network hubs [20]. Our findings in the current study support this hypothesis, as we have found that the relative hub-status of the RA varies largely within the patient cohort, and that whether it is more or less central than its neighbours does not determine outcome. Therefore, removal of a hub region was not necessary in this study to achieve seizure freedom. Of note, in this study we have considered the RA as a proxy for the epileptogenic zone, as it commonly done in epilepsy-surgery studies [22, 41, 43]. However, this adds a level of inaccuracy: for NSF cases it is known to be inaccurate, but even for SF cases it might have been larger that needed [20, 21]. This can lead to inaccuracies in the definition of the RA, and as a consequence of the neighbourhood regions. In contrast, the differences between either the RA or its neighbours with the remaining brain regions proved to be a stronger indicator of surgical outcome (albeit still weak, with *AUC* = 0.62 and 0.64, respectively). The proposed three-node-set partition may thus provide new insight into the effect of a particular resection, which may be missed with the standard two-node-set partition approach. This is in agreement with the methodology and findings in [34], but here we propose a methodology based only on resting-state MEG brain connectivity, without the need for invasive or ictal recordings, as the notion of the propagation zone is substituted by that of the neighbours of the RA.

### B. Centrality metrics and node neighbourhoods

In this study we proposed the use of regional centrality metrics to better account for the effect of a given resection, following previous theoretical works [56, 64, 65]. Most previous clinical studies have considered traditional centrality metrics that do not take the local network-neighbourhood into account, of which the degree [40–43], betweenness centrality [40, 41], and eigenvector centrality [20, 66, 67] are predominant. Here we found that neighbourhood-based metrics, with the exception of *β*_0_ (which equaled 1 in most cases for the considered parameters, as a consequence of the high level of recurrent connectivity in the networks), were able to more consistently pick up differences between ℛ𝒜 and 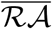 nodes at the individual level across all frequency bands, and in particular for the broadband, than nodal measures such as the betweenness centrality or the clustering coefficient (figure 3). These findings indicate that the neighbourhood of the RA is significantly different from the neighbourhood of other nodes in the brain network, in particular denoting a higher (generalized) centrality. In contrast, at the group level (figure 4) the metric that revealed the strongest difference between ℛ𝒜 and 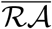 nodes was the betweenness centrality, which is also the metric most often considered in the literature. We note this as an interesting venue for future research: at the theoretical level to understand whether different centrality metrics might be more or less sensitive to individual variations, and at the clinical level to validate the generalizability of these findings. Notably, whereas the betweenness centrality requires of global information, extended neighbourhood metrics can be computed with only regional information, and are thus more efficient to compute for large systems.

In the case of the three-node-set partition, at the group-level the differences between local and regional centrality metrics were larger (figure 4 and Supp. Table 4). This may be caused by the neighbourhood-based partition of the node sets, such that the RA neighbourhood is considered explicitly, even for the node-based metrics. At the individual level the neighbourhood-based metrics were also slightly more sensitive to differences between both the RA (ℛ𝒜 set) and its neighbours (𝒩 set) with the remaining network nodes (𝒪 set). Differences between the ℛ𝒜 and 𝒩 node-sets were sparse as discussed above, and generally all metrics performed similarly except for the clustering coefficient *c*, and the first and third Betti numbers, *β*_0_ and *β*_2_, with very low sensitivity. In particular *β*_0_ and *β*_2_, showed little variation across nodes for the parameters considered. At the group level, however, the betweenness centrality and curvature found the strongest and more consistent differences between node-sets. Further studies, considering e.g., larger networks or different connectivity thresholds, could validate the generalizability of these findings.

In order to better contextualize our study, we also considered the node strength (or weighted degree) as a centrality metric, following [43]. In their original study the authors found that this metric could classify ℛ𝒜 and 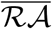 nodes for 8 out of 12 SF patients, and that, using the area under the curve of this classification (distinguishability *D*^*′*^) as a patient score, they could classify SF and NSF patients with an *AUC* of 0.76. In our study, however, we have only found an *AUC* of 0.65 when implementing their methodology, and an optimal value of *AUC* = 0.68 for the *α*_1_ band with the combined distinguishability score. These results are in agreement with those found in the main part of our study, and with our previous findings with this same dataset [41]. Further studies are needed to elucidate the origin of the lower performance found here compared to Ramaraju et al. We identify methodological considerations, such as the choice of connectivity metric –we considered here a phase metric, the PLI, that is insensitive to volume conduction, whereas Ramaraju et al. used uncorrected amplitude correlations [43]– or the thresholding procedure used (simple thresholding vs the disparity filter considered here). Moreover, the small dataset considered by Ramaraju et al. could have driven the higher performance of the classification analysis. The findings may also reflect intrinsic differences between the patient populations: the cohort in this study is highly heterogeneous, including patients with different etiologies.

### C. Multi-frequency analysis

In our study we considered a multi-band description, in analogy with some previous studies [35, 46, 62, 68–71]. These studies found for the most part comparable results across frequency bands, with significant differences in brain network organization between epilepsy patients and controls, or between SF and NSF epilepsy-surgery patients, arising predominantly in the *θ* and *α* bands [35, 46, 69–71], although differences have also been observed in the *δ* and *γ* bands [68] and in the ripple and fast ripple bands [68, 69].

In our study we also found comparable results across frequency bands for the node-based analyses, both at the individual and group level. Some metrics such as the local clustering *c, β*_0_ and *β*_2_ however only picked up differences between node sets in the broadband network. Notably, only in this band were the sizes of the 𝒩 and 𝒪 node-groups markedly different (when considering all ROIs and patients, see Supp. Figure 1 for more details). The bands for which we found the best patient classification were the broadband and *α*_1_, in agreement with the literature [35, 46, 69, 71]. Remarkably, we found the strongest variations across frequency bands in the patient classification analysis (figure 5). Whereas in the broadband and the lower frequency bands (in particular *δ* and *α*_1_) we found a somewhat better outcome for patients with high distinguishability score, this was not the case for higher frequency bands (in particular *β* and *γ*, see figure 5).

### D. Methodological considerations

In this study we considered the same patient database as in our previous study [41]. In this previous study, a machine learning analysis was used to classify network nodes as belonging or not to the resection area, and to classify patients as having good (SF) or bad (NSF) outcomes. The performance of the node classifier was fair (60.37% accuracy), but the patient classification failed (49.03% accuracy). We have introduced several methodological changes relative to this original study, from the consideration of multiple frequency bands, the three-node-group partition, and the inclusion of node-neighbourhoods and topological data analysis. The methodologies of the two studies can be compared via the betweenness centrality, a benchmark centrality measure considered in both studies: [41] found that hub nodes overlapped more than expected by chance with the ℛ𝒜. This is in qualitative agreement with our finding that ℛ𝒜 nodes are, at the group level, significantly more central than 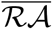 nodes.

Regarding the patient classification, [41] performed a classification based on a combination of individual and average metrics, namely the averages over (a) ℛ𝒜 nodes, (b) the resection lobe, (c) nodes contralateral to the ℛ𝒜, (d) 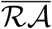 nodes, and two metrics measuring the difference between the average over ℛ𝒜 and the contralateral nodes, and over ℛ𝒜 and 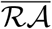 nodes. No significant differences between SF and NSF patients were identified at group level, and a machine learning analysis was also unable to classify the patients according to surgical outcome. In our study, instead of using the centrality values directly, we exploited the results of the node-based analyses to perform a patient classification analysis, similarly to [43]. In particular, we defined a distinguishability score based on the difference between each of the node sets, and we found an AUC of 0.68 for the broadband network (the same used in [41]). In this manner we were able to exploit a patient-specific analysis, accounting for heterogeneity in the patient population, which can be lost if comparisons of absolute values among patients are performed. The differences in findings between the two studies, and our finding that a population-based analysis is less sensitive than the patient-specific analysis, highlight the need to consider methodologies that allow for individualized patient characterization [22, 23].

Whereas some of the studies mentioned above [43], as well as other recent studies [22], have found better classification results than the ones found in this study, the strength of this study lies in the much larger patient cohort considered here, which is two to three times larger than typical cohort sizes in similar studies. Moreover, we further validated the robustness of our findings with respect to several methodological choices, including the frequency band of the MEG-based brain networks and specific analysis details, benchmarking our findings and analysis pipelines against previous studies [41, 43].

## IV. CONCLUSION

Pathological hubs occur in the brain networks of refractory-epilepsy patients that do not necessarily overlap with the epileptogenic zone, but may instead be strongly connected to it. Thus, a positive surgical outcome may be obtained also if the surgical resection does not include a pathological hub. In this study we have found that a three-group partition of the brain regions, where the neighbours of the resection area are separated from the remaining brain regions, can provide novel information regarding the organization of the epileptogenic network. Regional descriptors of hub-status and network organization, as the ones we propose here based on the notion of extended neighbourhoods, provide new tools to characterize the effect of a proposed resection. Our findings also evidence the heterogeneity of the patient population, and the need for individualized studies that allow for a patient-specific consideration of brain connectivity.

## V. METHODS

### A. Patient group

The patient cohort derived from the one presented in [41]. Three cases were removed, two due to existence of a previous resection, and one due to withdrawal of patient consent. The final patient cohort thus consisted of 91 patients with refractory epilepsy, with heterogeneous seizure etiology. All included patients (i) received a clinical MEG recording as part of their presurgical evaluation between 2010 and 2015 at Amsterdam University Medical Center, location VUmc; (ii) subsequently underwent epilepsy surgery at the same center; (iii) surgery outcome information was available following the Engel classification [9] either 1 year (88 patients) or at least 6 months (3 patients) after the surgery. A waiver of ethical review was obtained from the institutional review board (Medisch Ethische Toetsingscommissie Vrij Universiteit Medical Center) as no rules or procedures were imposed other than routine clinical care.

The patient group was heterogeneous with temporal and extratemporal resections and different etiology. Surgical outcome was classified according to the Engel classification [9]. 64 patients were deemed seizure free (SF).

### B. Individualized Brain Networks

Individualized brain networks were derived for each patient from 10 to 15 minute resting-state MEG (magnetoencephalography) recordings, using the Automated Anatomical Labeling (AAL) atlas [72] to define a brain parcellation of 90 Regions of Interest (ROIs), with 78 cortical and 12 subcortical ROIs, excluding the cereberallar ROIs [73]. The pre-processing steps, as well as the procedures to reconstruct the activity of each source are described in detail in [41]. We derived 7 brain networks for each patient: a broadband network (*B*, 0.5 − 48.0*Hz*) and six frequency-band specific networks: *δ* (0.5 − 4.0*Hz*), *θ* (4.0 − 8.0*Hz*), *α*_1_ (8.0 − 10.0*Hz*), *α*_2_ (10.0 − 12.0*Hz*), *β* (12.0 − 15.0*Hz*) and *γ* (15.0 − 30.0*Hz*), by filtering the source-reconstructed data in the corresponding frequency bands.

Each ROI defined one node in the network, and the coupling strength or link weight between each pair of nodes *w*_*ij*_ was estimated with the Phase Lag Index (PLI). The PLI is a functional connectivity metric that measures the asymmetry in the distribution of instantaneous phase differences between two times series [74]. The PLI is insensitive to zero-lag coupling and thus it is robust against volume conduction or field spread [74]. 174 epochs of 4096 samples (3.28s) where used for each patient to estimate functional coupling.

Raw PLI matrices were thresholded and binarized with a disparity filter method [75]. The disparity filter extracts the connectivity backbone (*a*_*ij*_ *>* 0 if there is a significant connection between *i* and *j* and 0 otherwise) of a network by removing connections that are not statistically significant. The disparity filter accounts for node heterogeneity in the edge weight distribution: weak edges are identified on a node-by-node basis, by comparing their strength to that of the remaining node’s edges with a given significance threshold *α* which we set to 0.1. This resulted in sparse networks (with network densities of about 5%; range: 0.047 − 0.051, see Supp. Table 1) with giant components spanning the majority of the nodes (range: 84.49 − 89.1).

### C. Local node metrics

We characterized the local structure of the network by three nodal properties. In particular, for each node *i* we considered its centrality (as given by the betweenness centrality *BC*_*i*_), clustering coefficient *CC*_*i*_, and curvature *C*_*i*_. The **betweenness centrality** of a node measures its influence over the flow of information on the graph: *BC*_*i*_ indicates the fraction of shortest paths in the network that pass through node *i*. The **local clustering coefficient** of a node indicates the fraction of triads involving node *i* that are closed, i.e.

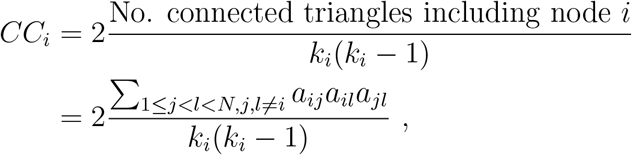

where *a*_*ij*_ indicates an element (*i, j*) of the adjacency matrix *A*, which is equal to 1 if *i* and *j* are connected by a link or 0 otherwise,*N* is the number of nodes in the network, and *k*_*i*_ = ∑_*j*_ *a*_*ij*_ is the degree of node *i*. Finally, the local curvature of a network generalizes the concept of curvature of a surface, which intuitively measures how the surface bends in distinct directions [76]:

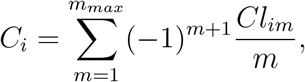

where *Cl*_*im*_ is the number of *m*-cliques see below to which *i* belongs, and *m*_*max*_ represents the size (i.e. number of nodes) of the largest clique in the network. Here we have considered the size of interactions up to three nodes (*m*_*max*_ = 3).

### D. Simplicial complex description

Simplicial complexes represent higher-order networks which allow for interaction between two but also more nodes, described by simplices. A *d*-simplex is formed by a set of *d* + 1 nodes and all their possible connections. For instance, a 0-simplex is simply a node, a 1-simplex a link and the two corresponding nodes, a 2-simplex is a triangle, a 3-simplex is a tetrahedron and so on. A simplicial complex 𝒦 is formed by a set of simplices such that i) if a simplex belongs to 𝒦 then any simplex formed by a subset of its nodes is also included in 𝒦, and ii) given two simplicies of 𝒦, their intersection either also belongs to 𝒦, or it is a null set [56]. A simplicial complex representation of a network can be built deterministically by defining the *clique complex* of the network. A *k*-clique is a subgraph of the network formed by *k* all-to-all connected nodes. That is, 1-cliques correspond to nodes, 2-cliques to links, 3-cliques to triangles, and so on. Thus, in order to build a simplicial complex of dimension *d* from a network, we identify all *d* + 1-cliques [56, 77]. This choice for creating simplices from cliques has the advantage of using pairwise signal processing to create a simplicial complex from brain networks [78]. Other strategies to build simplicial complexes beyond pairwise signal processing have been proposed, such as approaches combining information theory and algebraic topology [57, 79–83].

### E. Extended neighbourhood

The mesoscopic structure of a complex network can be described in terms of extended neighbourhoods or ego networks [56], as illustrated in figure 1 Starting from a given node *i*, we define its *d*−extended neighbourhood 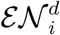 as the subgraph induced by the set of nodes at hopping distance *δ* equal or smaller to *d, δ* ≤ *d* (see figure 1b). 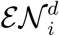 generalizes the concept of clustering coefficient, as it allows us to capture the connectivity not only between the first neighbours of a node, but of its general area of influence characterized by the hopping distance parameter *d*.

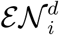 can be characterize by its size (number of nodes, 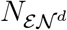) and connectivity (number of links, 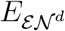). 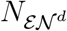 generalizes the notion of node degree, and indeed the degree of a node equals to 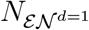. Similarly, the local clustering coefficient reduces to 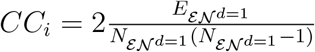.

Finally, we also characterized the topological organization of the extended neighbourhoods by the notion of *Betti numbers*. The first Betti number *β*_0_ measures the number of connected components on a network. Subsequent Betti numbers *β*_*i*_ describe the topology of the simplicial complex associated with the network. Generally, the Betti numbers *β*_*i*_, *i* ≥ 1 are topological invariants derived from the simplifical complex that measure the number of linearly independent *i*-dimensional holes in the simplicial complex. Thus, *β*_1_ provides the number of 1-dimensional cycles that are not boundaries of 2-dimensional simplices of the associated simplicial complex, and similarly *β*_2_ indicates the number of 2-dimensional cycles (i.e. over triangles) that are not boundaries of 3-dimensional simplices of the simplicial complex. *β*_0_ indicates the number of connected components of the local neighbourhood. Thus, large values indicate a hub that connects otherwise disconnected regions of the network [56]. *β*_1_ indicates the number of cycles forming 1-dimensional holes. Therefore, a large value of the ratio *β*_1_*/β*_0_ indicates a sparse neighbourhood. Similarly, larger values of *β*_2_ indicate the tendency to form planar (i.e. triangular) structures. The Betti numbers are non-linearly influenced by the size and density of the neighbourhood, and integrate information of the mesoscopic structure of the network in a non-trivial manner.

### F. Resection area and node sets

The resection area was determined for each patient from the three-month post-operative MRI. This was co-registered to the pre-operative MRI (used for the MEG co-registration) using FSL FLIRT (version 4.1.6) 12 parameter affine transformation. The resection area was then visually identified and assigned to the corresponding AAL ROIs, namely those for which at least 50% or the centroid had been removed during surgery.

Based on the resection area, we identified four sets of nodes: ℛ𝒜, or resected nodes, are the nodes that belong to the resection area. 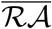, or non-resected nodes, are the nodes that do not belong to ℛ𝒜. We further considered two subsets of 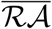 nodes. This partition was based on the connectivity of the resection area, and was thus different for each frequency band: 𝒩, or neighbours, are the nodes that are connected to ℛ𝒜 nodes and that do not themselves belong to the resection area. 𝒪, or other nodes, are the remaining nodes in the network, that is, nodes that do not belong to the resection area and are not connected to any ℛ𝒜 nodes.

### G. Statistical analyses

We first performed an individualized node-based analysis by which we tested whether the hub-status of the different node-sets differed significantly for each patient and metric 𝒳. We considered two types of comparisons: a) two-node-set setting, where we tested whether 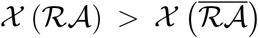, and b) three-node-set setting, where we tested whether 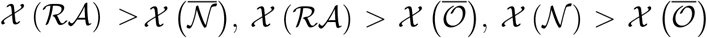. We quantified whether the hubness distributions were significantly different via bootstrapping analyses with 10^4^ replicas to determine the *z*-score and *p*-value of the difference. The sign of the difference indicated whether it was in the direction of the hypothesis or against it. The *z*-score was computed as the mean of the differences of the bootstrapped samples divided by the standard deviation of these differences. The 2-tailed *p*-value associated with the *z*-score was determined using the cumulative distribution function of a standard normal distribution. Considering the large number of comparisons performed, we applied the Bonferroni correction to account for multiple testing and control the false discovery rate. Specifically, the Bonferroni correction was applied by dividing the original significance level (*α* = 0.05) by the number of comparisons made. For each pairwise comparison of node-sets, we conducted 56 statistical comparisons (7 bands times 8 metrics). Thus, for every pair of nodes, we used the Bonferroni correction by adjusting the significance level to 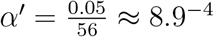.

To determine whether the results held at the group level, for each patient we estimated the average of each hubness metric for each of the node sets. We then performed a paired bootstrapping analysis to test whether the distribution of average metrics was significantly different, for each of the four pairs of node-comparisons as defined above.

We subsequently utilized the results of the node-based analyses to perform a receiver operating characteristic (ROC) curve classification of the patients (SF or NSF). The result of each node-based test was quantified in the variable 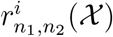 for each patient *i*, hubness metric 𝒳, and node-set sets *n*_1_ and 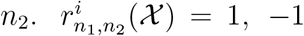 or 0 indicating whether the node-sets were significantly different in the direction of the hypothesis, contrary to it, or not significantly different, respectively. We then summed over hubness metrics to define a *distinguishability* score 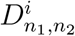 for each patient and node-based comparison [43]. To sum up the results of the three-node-set analysis, we defined a combined distinguishability score, 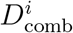, by summing over the corresponding three pairwise comparisons. The distinguishability score according to each test was then used to classify the patients with a receiver operating characteristic (ROC) curve analysis, and the goodness of the classification was measured with the area under the curve (AUC).

Finally, to enable a more direct comparison with the previous study by [43], we also estimated the distinguishability score as originally proposed by calculating the AUC of the node ROC-classification (instead of using 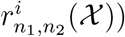, for each metric *X* and pair of nodesets. Patient-classification based on this score was then performed similarly to the previous analysis. For this test we also considered the node strength (the sum of its non-zero weights after thresholding) as a metric to allow for a more direct comparison with [43].

### H. Data availability

The data used for this manuscript are not publicly available because the patients did not consent for the sharing of their clinically obtained data. Requests for access to the data should be directed to the corresponding author. All user-developed codes are available on github: https://github.com/LeonardoDiGaetano/TDA-Epilepsy.

## Data Availability

The data used for this manuscript are not publicly available because the patients did not consent for the sharing of their clinically obtained data.
Requests for access to the data should be directed to the corresponding author.

## ACKNOWLEDGMENTS

A.P.M. acknowledges financial support by the “Ramón y Cajal” program of the Spanish Ministry of Science and Innovation (grant RYC2021-031241-I), and from the Spanish Research Agency, Project PID2020-113681GBI00, financed by MICIN/AEI/10.13039/501100011033. A.P.M. and I.N. were supported by ZonMw and the Dutch Epilepsy Foundation, project number 95105006, and Dutch Epilepsy Foundation project 14–16 (I.N.) L.D.G. acknowledges financial support from the Clinical Neurophysiology Department (KNF) at the Amstedam University Medical Center (AUMC). The funding sources had no role in study design, data collection and analysis, interpretation of results, decision to publish, or preparation of the manuscript. N.D. acknowledges funding by the Swiss National Science Foundation (SNSF) under project funding ID: 200021 207537 and by the Deutsche Forschungsgemeinschaft (DFG, German Research Foundation) under Germany’s Excellence Strategy EXC2181/1-390900948 (the Heidelberg STRUCTURES Excellence Cluster). This work was partially supported by a grant from the Simons Foundation (G. B.). G.B. and A. P. M. would like to thank the Isaac Newton Institute for Mathematical Sciences, Cambridge, for support and hospitality during the programme Hypergraphs: Theory and Applications, where work on this paper was undertaken. This work was supported by EPSRC grant EP/R014604/1.

## VI. COMPETING INTERESTS

The authors declare that they have no competing interests.

All authors participated in the study design. L.D.G., F.A.N.S. and A.P.M. devised the computational and visualization methods. L.D.G. and A.P.M. performed the analyses. L.D.G. and A.P.M. wrote the original draft and all authors participated in writing, review and editing.

## SUPPLEMENTARY INFORMATION

### S.1. DISPARITY FILTER: BASIC NETWORK STATISTICS

The brain networks were thresholded with a disparity filter with significance threshold of *α* = 0.1. In Supp. Table S.1 we report the average number of edges remaining in the network after the thresholding procedure, and the size of the giant component.

**TABLE S.1:**
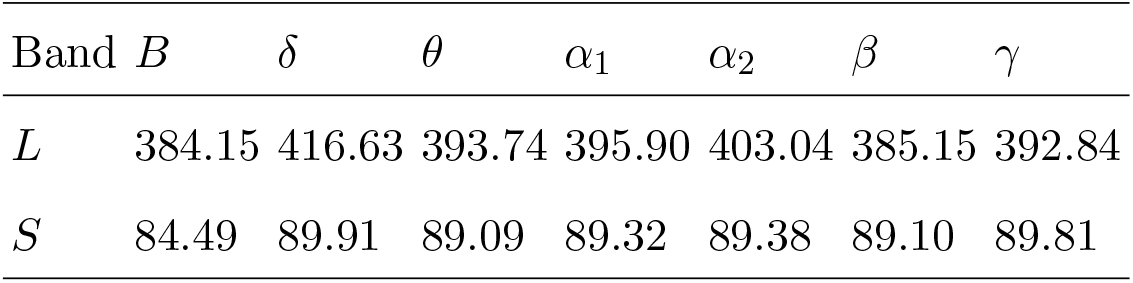
Basic network statistics. Average number of edges *L* remaining in the network after the thresholding procedure, and the average size of the largest component *S*, for each frequency band.

### S.2. BASIC NODE-SETS STATISTICS

In figure S.1 we report the distribution of node-set sizes for each frequency band.

**FIG. S.1:**
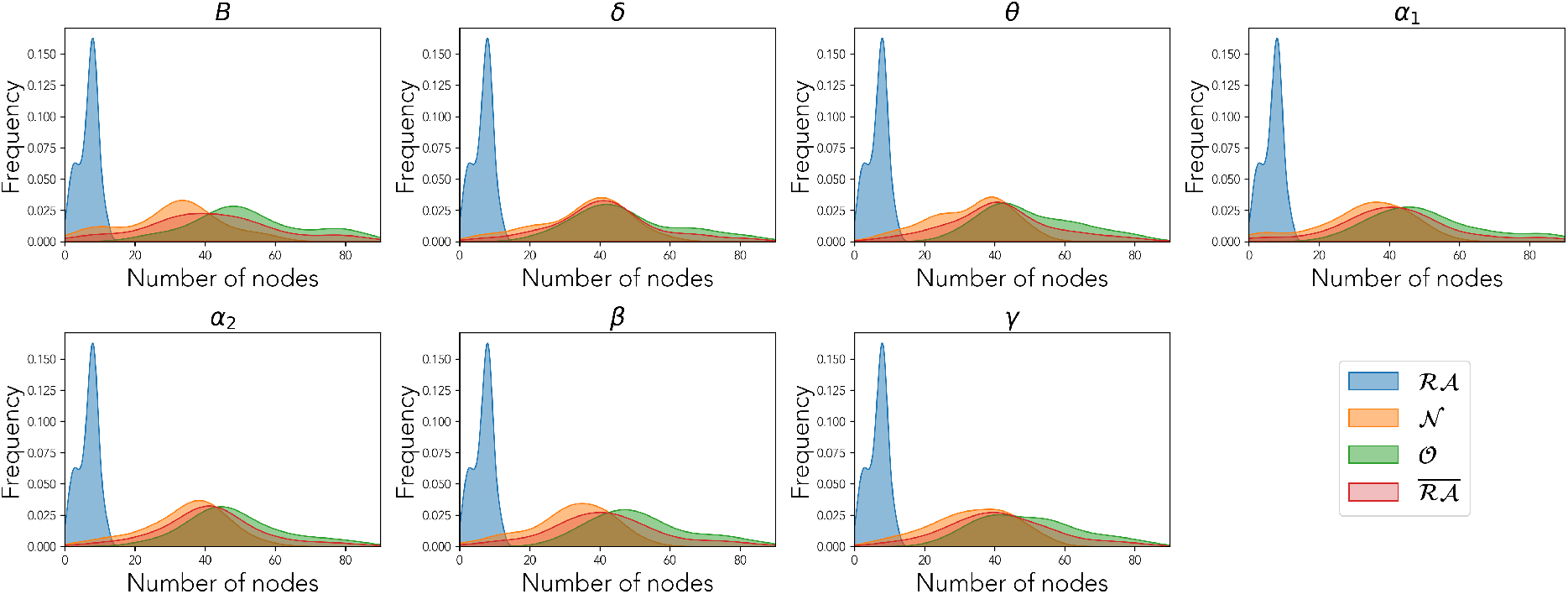
Distributions of the size of each node-set (as indicated by the general legend) over the patient population, for each frequency band as indicated by the panel title.

### S.3. METRICS OF NETWORK TOPOLOGY

To quantify the connectivity and network properties of brain nodes, we utilized a variety of metrics. These metrics can be categorized into node-level metrics and neighbourhood-level metrics. Below is a detailed description of each metric:

#### A. Node-level Metrics

- The **Betweenness Centrality** *BC* measures the influence of a node over the flow of information within the network. It is calculated by determining the fraction of all shortest paths in the network that pass through a given node. Nodes with high betweenness centrality are considered critical for information transfer and can be identified as hubs within the network [58].
- The **Local Clustering Coefficient** *c* quantifies the extent to which nodes in a graph tend to form clusters or groups. For a given node *i*, the clustering coefficient is defined as the ratio of the number of closed triplets (or triangles) to the total number of triplets (both open and closed) centered on that node. Mathematically, it is given by:

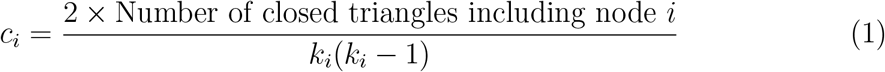

where *k*_*i*_ is the degree of node *i*. A higher clustering coefficient indicates a greater tendency for node *i* to form tightly-knit groups with its neighbors [58].
- The **Local Curvature** *C* captures how paths bend around a node in its vicinity, offering insights into the local geometric structure More specifically:

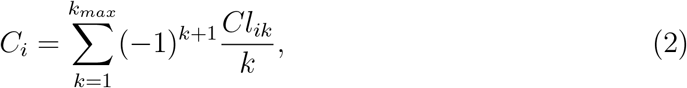

where *Cl*_*ik*_ is the number of *k*-cliques to which *i* belongs, and *k*_*max*_ represents the size (i.e. number of nodes) of the largest clique in the network (*k*_*max*_ = 3 considering interactions up to three-node ones.). It generalizes the concept of curvature from differential geometry to network theory. Nodes with high curvature tend to have a significant influence on the robustness and stability of the network [76].

#### B. Neighbourhood-level Metrics

The extended neighbourhood *ε*𝒩 of a node encompasses all nodes within a certain distance (or hops) from the given node, excluding the node itself [56].

- The **Number of Nodes in the Extended Neighbourhood** *N* measures the size of the *ε*𝒩 and it generalizes the concept of node degree.
- The **Number of Edges in the Extended Neighbourhood** *E* quantifies the total number of pairwise edges within the *ε*𝒩, reflecting the local connectivity density.
- The **Betti Numbers (***β*_0_, *β*_1_, *β*_2_) are topological invariants that describe the connectivity of simplicial complexes (contructed in this case from the node neighbourhoods) at different dimensions, generalizing the notion of clustering coefficient:
  - *β*_0_ represents the number of connected components in the *ε*𝒩, indicating the degree of fragmentation. A higher *β*_0_ indicates a node that acts as a broker between different communities.
  - *β*_1_ quantifies the number of one-dimensional holes or open loops representing independent cycles within the *ε*𝒩. It provides information on the presence of circular structures that are not filled in by higher-dimensional simplices.
  - *β*_2_ measures the number of two-dimensional voids, reflecting higher-order connectivity patterns such as cavities within the *ε*𝒩.

In the main section of this paper we used these 8 metrics to describe local and regional network organization for each node, for each patient- and frequency-specific brain network. In Supp. Figure S.2 we show an illustrative example of the distribution of values for each of these metrics, for each of the node-sets defined in the main text, namely nodes in and outside the resection area (ℛ𝒜 and 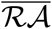 node sets), the neighbours of the resection area 𝒩 and other nodes in the network 𝒪.

**FIG. S.2:**
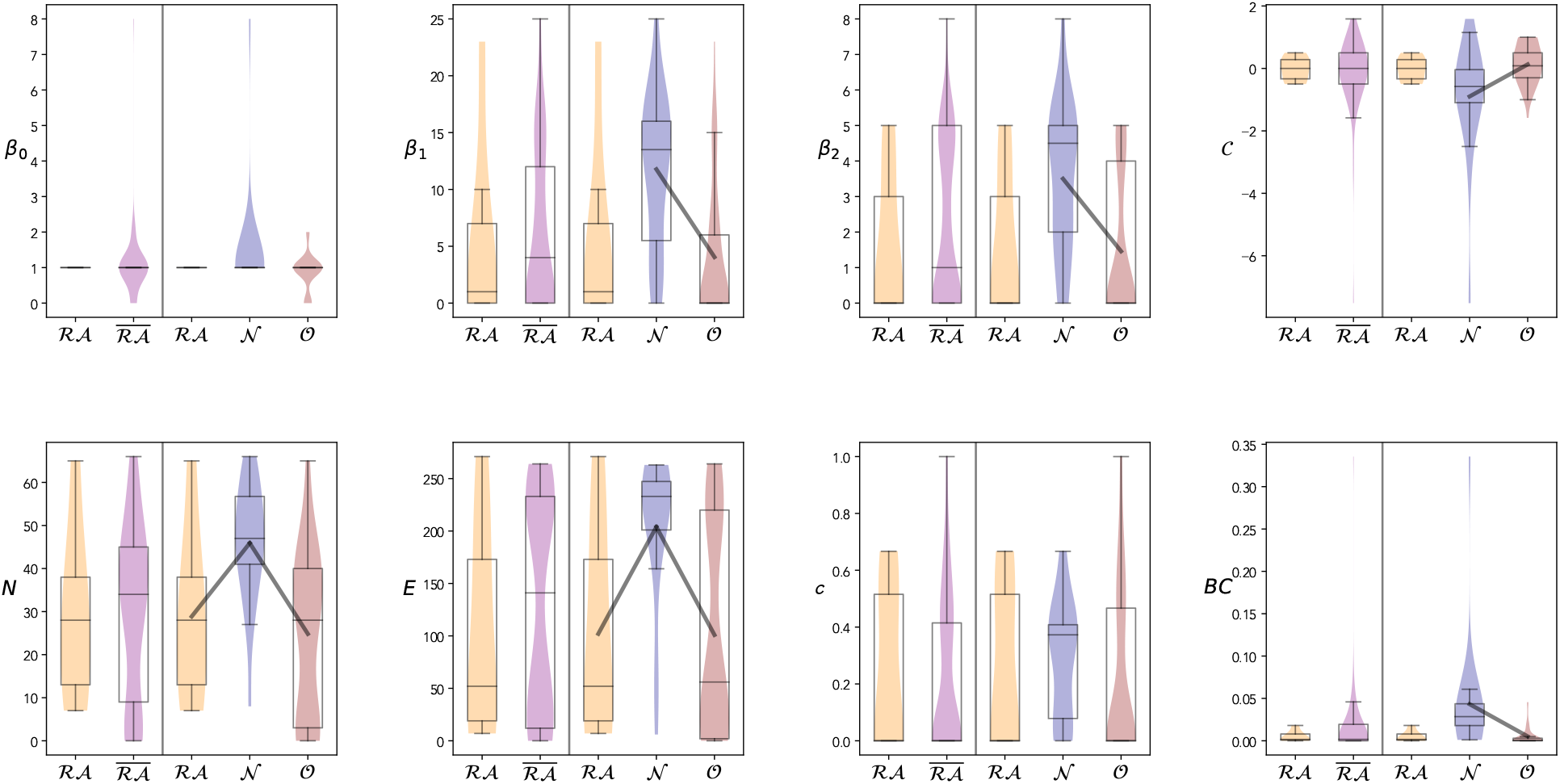
Distribution of generalized centrality metrics for an exemplary case (SF patient, broadband network) for each node-set. Each panel corresponds to a generalized centrality metric as indicated by the labels. For each panel we show the results for the two analysis that were performed: the two-node-set partition (left) accounting for the (ℛ𝒜 and 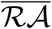 node sets, and the three-node-set partition (right) accounting for the (ℛ𝒜, 𝒩 and 𝒪 node sets. In all panels we show the distribution of values for each node-set as a violin plot, and indicate the mean and median values with solid lines. The box-plots indicate the median, the 25% and 75% percentiles and the extreme values. Significant differences between two groups are indicated by black lines connecting the corresponding violins.

### S.4. SUPPLEMENTARY INFORMATION TO PATIENT-SPECIFIC COMPARISON

Here we report the number of patients for whom there was a significant difference in the direction of the hypothesis (Table S.2) and in opposite direction (Table S.3) relative to the results presented in figure 3 of the main material.

**TABLE S.2:**
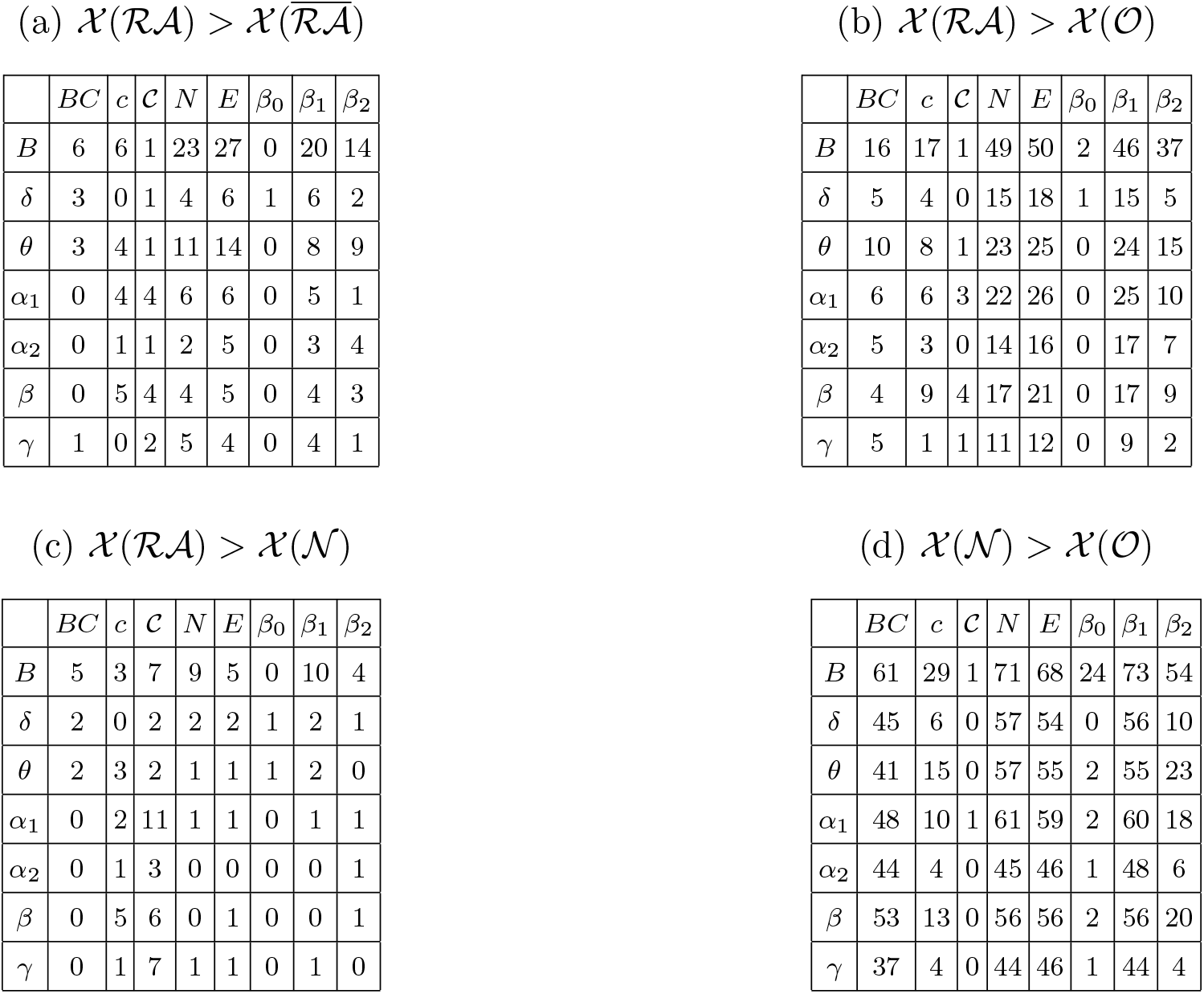
Number of patients for whom there was a significant difference in the direction of the hypothesis of figure 3 of the main text.

| | $BC$ | $c$ | $C$ | $N$ | $E$ | $\beta_0$ | $\beta_1$ | $\beta_2$ |
| --- | --- | --- | --- | --- | --- | --- | --- | --- |
| $B$ | 6 | 6 | 1 | 23 | 27 | 0 | 20 | 14 |
| $\delta$ | 3 | 0 | 1 | 4 | 6 | 1 | 6 | 2 |
| $\theta$ | 3 | 4 | 1 | 11 | 14 | 0 | 8 | 9 |
| $\alpha_1$ | 0 | 4 | 4 | 6 | 6 | 0 | 5 | 1 |
| $\alpha_2$ | 0 | 1 | 1 | 2 | 5 | 0 | 3 | 4 |
| $\beta$ | 0 | 5 | 4 | 4 | 5 | 0 | 4 | 3 |
| $\gamma$ | 1 | 0 | 2 | 5 | 4 | 0 | 4 | 1 |

(b) $\mathcal{X}(\mathcal{RA}) > \mathcal{X}(\mathcal{O})$
| | $BC$ | $c$ | $C$ | $N$ | $E$ | $\beta_0$ | $\beta_1$ | $\beta_2$ |
| --- | --- | --- | --- | --- | --- | --- | --- | --- |
| $B$ | 16 | 17 | 1 | 49 | 50 | 2 | 46 | 37 |
| $\delta$ | 5 | 4 | 0 | 15 | 18 | 1 | 15 | 5 |
| $\theta$ | 10 | 8 | 1 | 23 | 25 | 0 | 24 | 15 |
| $\alpha_1$ | 6 | 6 | 3 | 22 | 26 | 0 | 25 | 10 |
| $\alpha_2$ | 5 | 3 | 0 | 14 | 16 | 0 | 17 | 7 |
| $\beta$ | 4 | 9 | 4 | 17 | 21 | 0 | 17 | 9 |
| $\gamma$ | 5 | 1 | 1 | 11 | 12 | 0 | 9 | 2 |

(c) $\mathcal{X}(\mathcal{RA}) > \mathcal{X}(\mathcal{N})$
| | $BC$ | $c$ | $C$ | $N$ | $E$ | $\beta_0$ | $\beta_1$ | $\beta_2$ |
| --- | --- | --- | --- | --- | --- | --- | --- | --- |
| $B$ | 5 | 3 | 7 | 9 | 5 | 0 | 10 | 4 |
| $\delta$ | 2 | 0 | 2 | 2 | 2 | 1 | 2 | 1 |
| $\theta$ | 2 | 3 | 2 | 1 | 1 | 1 | 2 | 0 |
| $\alpha_1$ | 0 | 2 | 11 | 1 | 1 | 0 | 1 | 1 |
| $\alpha_2$ | 0 | 1 | 3 | 0 | 0 | 0 | 0 | 1 |
| $\beta$ | 0 | 5 | 6 | 0 | 1 | 0 | 0 | 1 |
| $\gamma$ | 0 | 1 | 7 | 1 | 1 | 0 | 1 | 0 |

| | $BC$ | $c$ | $C$ | $N$ | $E$ | $\beta_0$ | $\beta_1$ | $\beta_2$ |
| --- | --- | --- | --- | --- | --- | --- | --- | --- |
| $B$ | 61 | 29 | 1 | 71 | 68 | 24 | 73 | 54 |
| $\delta$ | 45 | 6 | 0 | 57 | 54 | 0 | 56 | 10 |
| $\theta$ | 41 | 15 | 0 | 57 | 55 | 2 | 55 | 23 |
| $\alpha_1$ | 48 | 10 | 1 | 61 | 59 | 2 | 60 | 18 |
| $\alpha_2$ | 44 | 4 | 0 | 45 | 46 | 1 | 48 | 6 |
| $\beta$ | 53 | 13 | 0 | 56 | 56 | 2 | 56 | 20 |
| $\gamma$ | 37 | 4 | 0 | 44 | 46 | 1 | 44 | 4 |

(a) $\mathcal{X}(\mathcal{RA}) > \mathcal{X}(\overline{\mathcal{RA}})$

**TABLE S.3:**
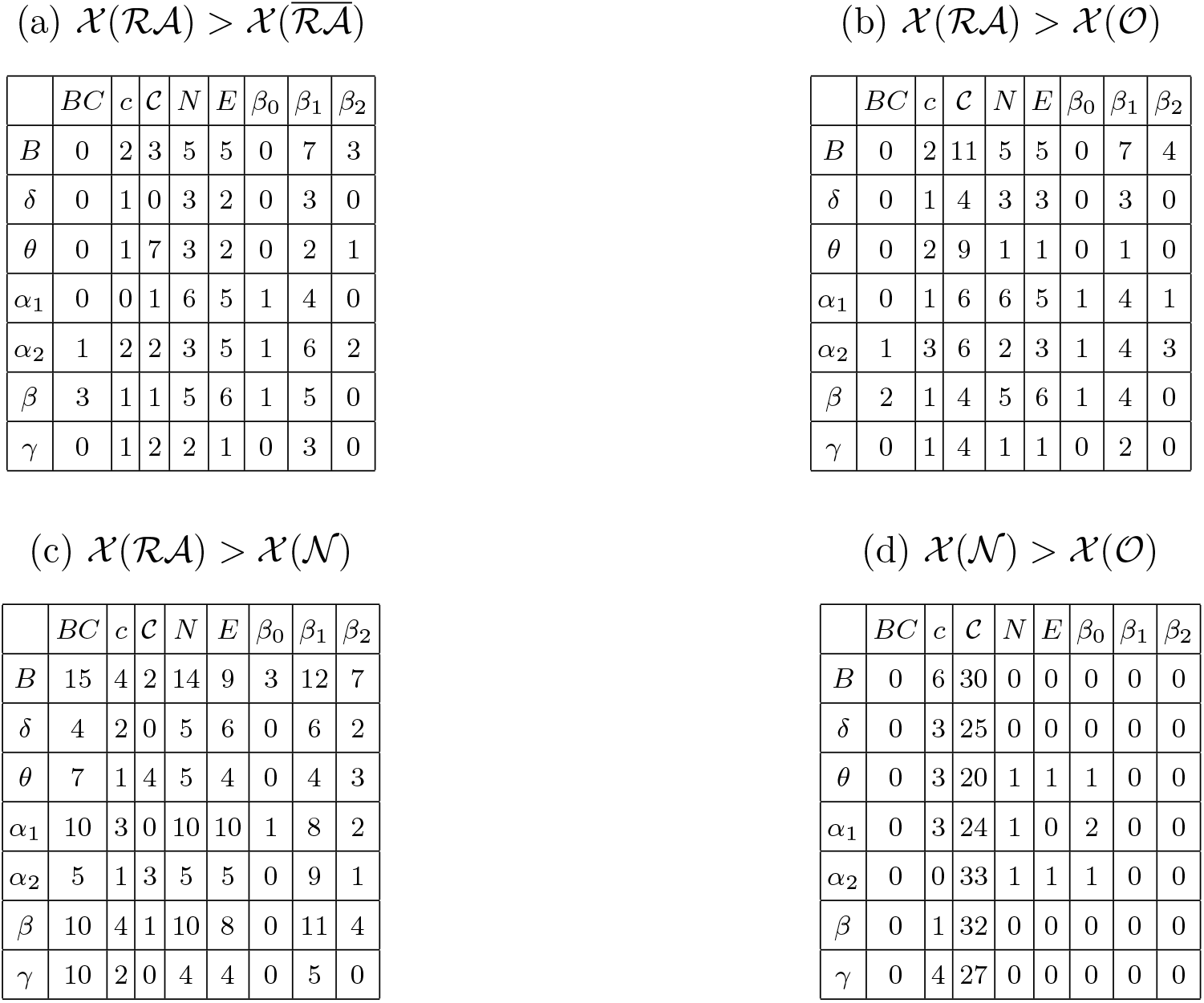
Number of patients for whom there was a significant difference in opposite direction of the hypothesis of figure 3 of the main text.

| | $BC$ | $c$ | $C$ | $N$ | $E$ | $\beta_0$ | $\beta_1$ | $\beta_2$ |
| --- | --- | --- | --- | --- | --- | --- | --- | --- |
| $B$ | 0 | 2 | 3 | 5 | 5 | 0 | 7 | 3 |
| $\delta$ | 0 | 1 | 0 | 3 | 2 | 0 | 3 | 0 |
| $\theta$ | 0 | 1 | 7 | 3 | 2 | 0 | 2 | 1 |
| $\alpha_1$ | 0 | 0 | 1 | 6 | 5 | 1 | 4 | 0 |
| $\alpha_2$ | 1 | 2 | 2 | 3 | 5 | 1 | 6 | 2 |
| $\beta$ | 3 | 1 | 1 | 5 | 6 | 1 | 5 | 0 |
| $\gamma$ | 0 | 1 | 2 | 2 | 1 | 0 | 3 | 0 |

(b) $\mathcal{X}(\mathcal{RA}) > \mathcal{X}(\mathcal{O})$
| | $BC$ | $c$ | $C$ | $N$ | $E$ | $\beta_0$ | $\beta_1$ | $\beta_2$ |
| --- | --- | --- | --- | --- | --- | --- | --- | --- |
| $B$ | 0 | 2 | 11 | 5 | 5 | 0 | 7 | 4 |
| $\delta$ | 0 | 1 | 4 | 3 | 3 | 0 | 3 | 0 |
| $\theta$ | 0 | 2 | 9 | 1 | 1 | 0 | 1 | 0 |
| $\alpha_1$ | 0 | 1 | 6 | 6 | 5 | 1 | 4 | 1 |
| $\alpha_2$ | 1 | 3 | 6 | 2 | 3 | 1 | 4 | 3 |
| $\beta$ | 2 | 1 | 4 | 5 | 6 | 1 | 4 | 0 |
| $\gamma$ | 0 | 1 | 4 | 1 | 1 | 0 | 2 | 0 |

(c) $\mathcal{X}(\mathcal{RA}) > \mathcal{X}(\mathcal{N})$
| | $BC$ | $c$ | $C$ | $N$ | $E$ | $\beta_0$ | $\beta_1$ | $\beta_2$ |
| --- | --- | --- | --- | --- | --- | --- | --- | --- |
| $B$ | 15 | 4 | 2 | 14 | 9 | 3 | 12 | 7 |
| $\delta$ | 4 | 2 | 0 | 5 | 6 | 0 | 6 | 2 |
| $\theta$ | 7 | 1 | 4 | 5 | 4 | 0 | 4 | 3 |
| $\alpha_1$ | 10 | 3 | 0 | 10 | 10 | 1 | 8 | 2 |
| $\alpha_2$ | 5 | 1 | 3 | 5 | 5 | 0 | 9 | 1 |
| $\beta$ | 10 | 4 | 1 | 10 | 8 | 0 | 11 | 4 |
| $\gamma$ | 10 | 2 | 0 | 4 | 4 | 0 | 5 | 0 |

| | $BC$ | $c$ | $C$ | $N$ | $E$ | $\beta_0$ | $\beta_1$ | $\beta_2$ |
| --- | --- | --- | --- | --- | --- | --- | --- | --- |
| $B$ | 0 | 6 | 30 | 0 | 0 | 0 | 0 | 0 |
| $\delta$ | 0 | 3 | 25 | 0 | 0 | 0 | 0 | 0 |
| $\theta$ | 0 | 3 | 20 | 1 | 1 | 1 | 0 | 0 |
| $\alpha_1$ | 0 | 3 | 24 | 1 | 0 | 2 | 0 | 0 |
| $\alpha_2$ | 0 | 0 | 33 | 1 | 1 | 1 | 0 | 0 |
| $\beta$ | 0 | 1 | 32 | 0 | 0 | 0 | 0 | 0 |
| $\gamma$ | 0 | 4 | 27 | 0 | 0 | 0 | 0 | 0 |

### S.5. SUPPLEMENTARY INFORMATION TO GROUP-LEVEL COMPARISON

Here we report the numerical values presented in figure 4 of the main material. Table S.4 presents z-scores presented through colors in figure 4 of the main material and Table S.5 the corresponding p-values.

**TABLE S.4:**
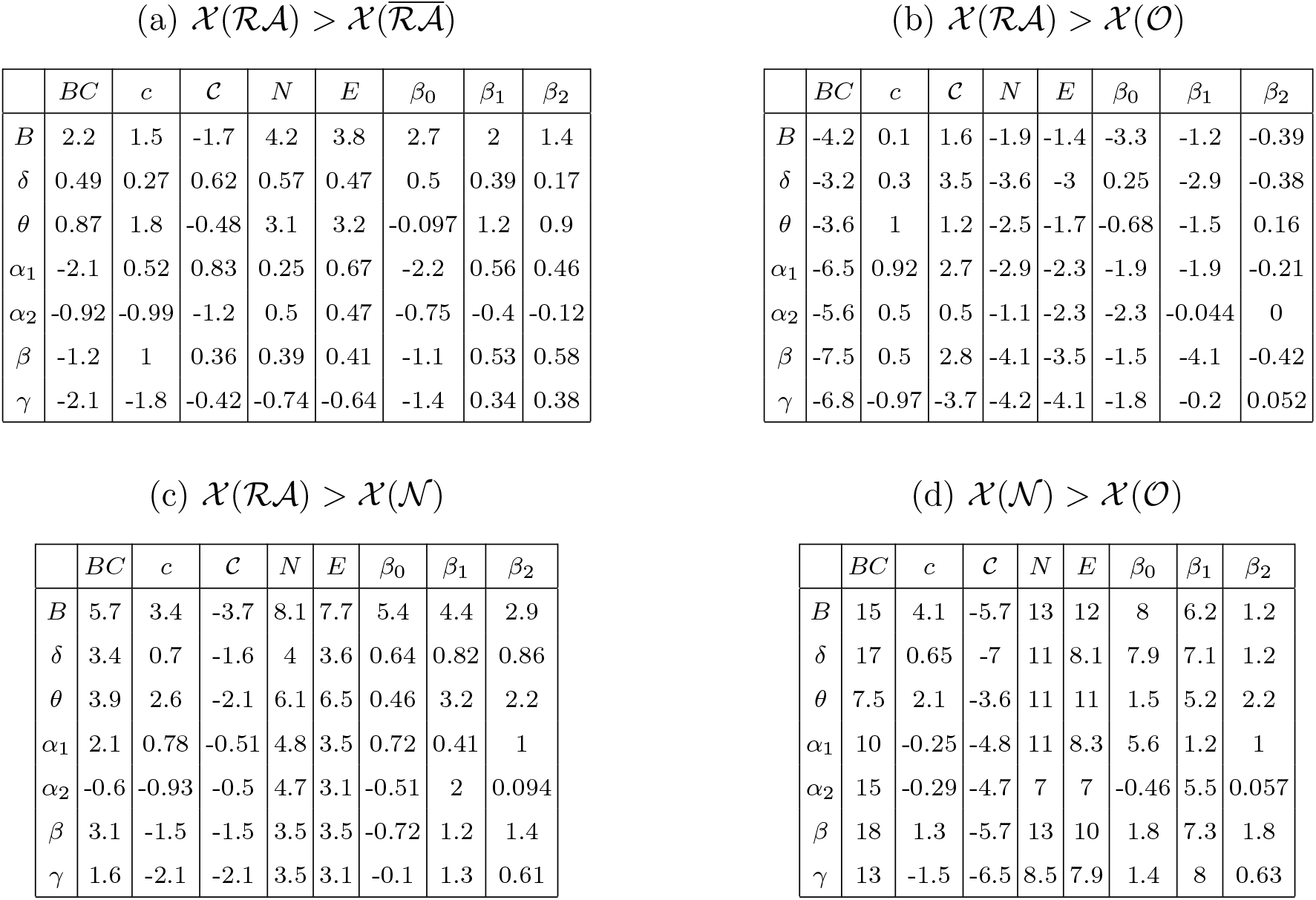
Numerical values corresponding to results of figure 4 of the main text. Group-level comparison between nodes sets, for each considered frequency-band (y-axis) and network metric (x-axis). 𝒳(*S*) stands for the generalized centrality metric 𝒳 measured on the nodes in set *S*. The numbers indicate the *z*-score of the difference between the average values of each node set, computed by bootstrapping the data (sampling size of 10^4^).

| | $BC$ | $c$ | $\mathcal{C}$ | $N$ | $E$ | $\beta_0$ | $\beta_1$ | $\beta_2$ |
| --- | --- | --- | --- | --- | --- | --- | --- | --- |
| $B$ | 2.2 | 1.5 | -1.7 | 4.2 | 3.8 | 2.7 | 2 | 1.4 |
| $\delta$ | 0.49 | 0.27 | 0.62 | 0.57 | 0.47 | 0.5 | 0.39 | 0.17 |
| $\theta$ | 0.87 | 1.8 | -0.48 | 3.1 | 3.2 | -0.097 | 1.2 | 0.9 |
| $\alpha_1$ | -2.1 | 0.52 | 0.83 | 0.25 | 0.67 | -2.2 | 0.56 | 0.46 |
| $\alpha_2$ | -0.92 | -0.99 | -1.2 | 0.5 | 0.47 | -0.75 | -0.4 | -0.12 |
| $\beta$ | -1.2 | 1 | 0.36 | 0.39 | 0.41 | -1.1 | 0.53 | 0.58 |
| $\gamma$ | -2.1 | -1.8 | -0.42 | -0.74 | -0.64 | -1.4 | 0.34 | 0.38 |

(b) $\mathcal{X}(\mathcal{RA}) > \mathcal{X}(\mathcal{O})$
| | $BC$ | $c$ | $\mathcal{C}$ | $N$ | $E$ | $\beta_0$ | $\beta_1$ | $\beta_2$ |
| --- | --- | --- | --- | --- | --- | --- | --- | --- |
| $B$ | -4.2 | 0.1 | 1.6 | -1.9 | -1.4 | -3.3 | -1.2 | -0.39 |
| $\delta$ | -3.2 | 0.3 | 3.5 | -3.6 | -3 | 0.25 | -2.9 | -0.38 |
| $\theta$ | -3.6 | 1 | 1.2 | -2.5 | -1.7 | -0.68 | -1.5 | 0.16 |
| $\alpha_1$ | -6.5 | 0.92 | 2.7 | -2.9 | -2.3 | -1.9 | -1.9 | -0.21 |
| $\alpha_2$ | -5.6 | 0.5 | 0.5 | -1.1 | -2.3 | -2.3 | -0.044 | 0 |
| $\beta$ | -7.5 | 0.5 | 2.8 | -4.1 | -3.5 | -1.5 | -4.1 | -0.42 |
| $\gamma$ | -6.8 | -0.97 | -3.7 | -4.2 | -4.1 | -1.8 | -0.2 | 0.052 |

(c) $\mathcal{X}(\mathcal{RA}) > \mathcal{X}(\mathcal{N})$
| | $BC$ | $c$ | $\mathcal{C}$ | $N$ | $E$ | $\beta_0$ | $\beta_1$ | $\beta_2$ |
| --- | --- | --- | --- | --- | --- | --- | --- | --- |
| $B$ | 5.7 | 3.4 | -3.7 | 8.1 | 7.7 | 5.4 | 4.4 | 2.9 |
| $\delta$ | 3.4 | 0.7 | -1.6 | 4 | 3.6 | 0.64 | 0.82 | 0.86 |
| $\theta$ | 3.9 | 2.6 | -2.1 | 6.1 | 6.5 | 0.46 | 3.2 | 2.2 |
| $\alpha_1$ | 2.1 | 0.78 | -0.51 | 4.8 | 3.5 | 0.72 | 0.41 | 1 |
| $\alpha_2$ | -0.6 | -0.93 | -0.5 | 4.7 | 3.1 | -0.51 | 2 | 0.094 |
| $\beta$ | 3.1 | -1.5 | -1.5 | 3.5 | 3.5 | -0.72 | 1.2 | 1.4 |
| $\gamma$ | 1.6 | -2.1 | -2.1 | 3.5 | 3.1 | -0.1 | 1.3 | 0.61 |

TABLE S.4: Numerical values corresponding to results of figure 4 of the main text.
| | $BC$ | $c$ | $\mathcal{C}$ | $N$ | $E$ | $\beta_0$ | $\beta_1$ | $\beta_2$ |
| --- | --- | --- | --- | --- | --- | --- | --- | --- |
| $B$ | 15 | 4.1 | -5.7 | 13 | 12 | 8 | 6.2 | 1.2 |
| $\delta$ | 17 | 0.65 | -7 | 11 | 8.1 | 7.9 | 7.1 | 1.2 |
| $\theta$ | 7.5 | 2.1 | -3.6 | 11 | 11 | 1.5 | 5.2 | 2.2 |
| $\alpha_1$ | 10 | -0.25 | -4.8 | 11 | 8.3 | 5.6 | 1.2 | 1 |
| $\alpha_2$ | 15 | -0.29 | -4.7 | 7 | 7 | -0.46 | 5.5 | 0.057 |
| $\beta$ | 18 | 1.3 | -5.7 | 13 | 10 | 1.8 | 7.3 | 1.8 |
| $\gamma$ | 13 | -1.5 | -6.5 | 8.5 | 7.9 | 1.4 | 8 | 0.63 |

(a) $\mathcal{X}(\mathcal{RA}) > \mathcal{X}(\overline{\mathcal{RA}})$

**TABLE S.5:**
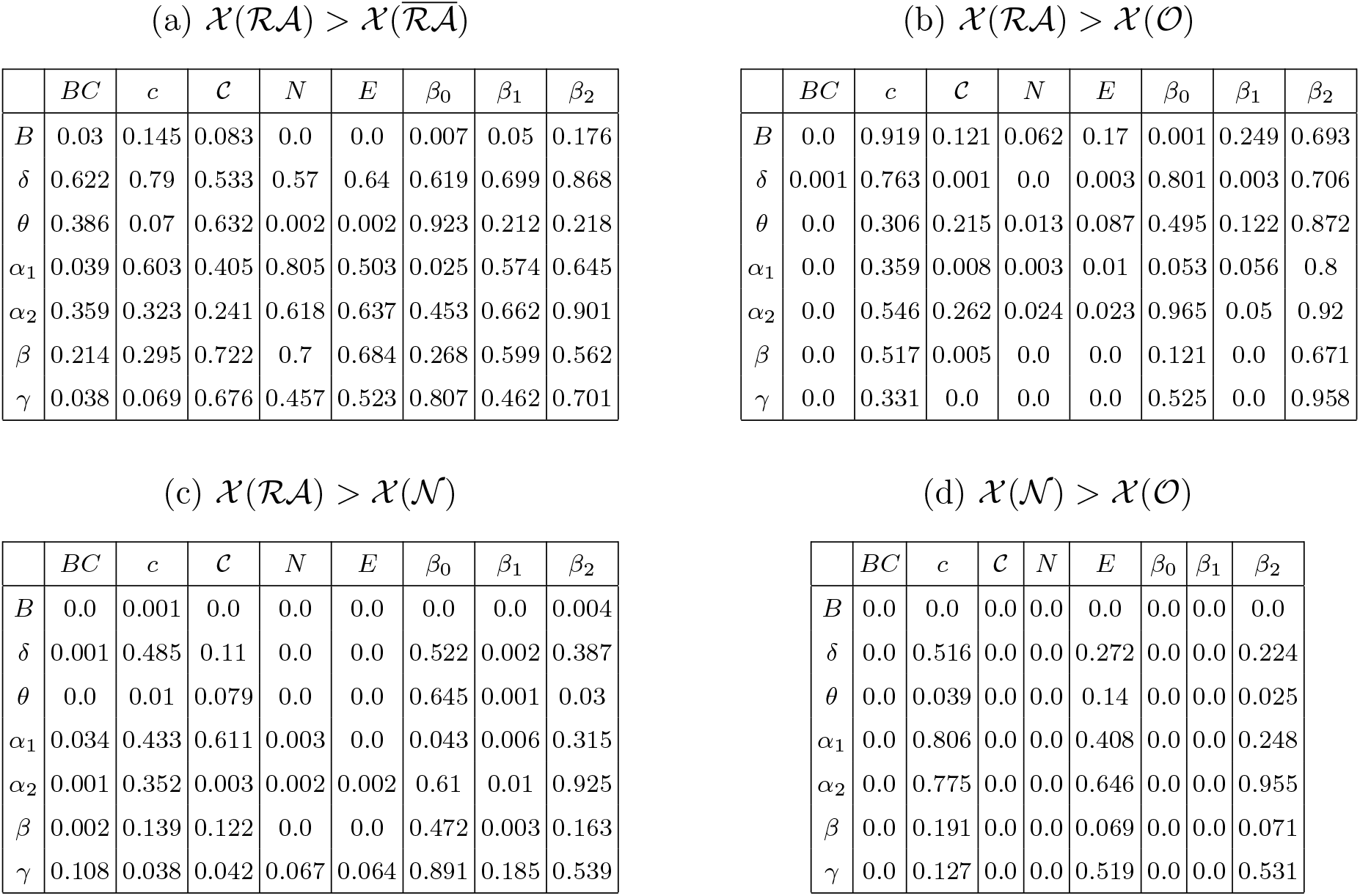
p-values corresponding to results of figure 4 of the main text. Group-level comparison between nodes sets, for each considered frequency-band (y-axis) and network metric (x-axis). 𝒳 (*S*) stands for the generalized centrality metric 𝒳 measured on the nodes in set *S*. The numbers indicate the p-values of the comparison between the average values of each node set presented in Table S.4 and figure 4 of the main text.

| | $BC$ | $c$ | $\mathcal{C}$ | $N$ | $E$ | $\beta_0$ | $\beta_1$ | $\beta_2$ |
| --- | --- | --- | --- | --- | --- | --- | --- | --- |
| $B$ | 0.03 | 0.145 | 0.083 | 0.0 | 0.0 | 0.007 | 0.05 | 0.176 |
| $\delta$ | 0.622 | 0.79 | 0.533 | 0.57 | 0.64 | 0.619 | 0.699 | 0.868 |
| $\theta$ | 0.386 | 0.07 | 0.632 | 0.002 | 0.002 | 0.923 | 0.212 | 0.218 |
| $\alpha_1$ | 0.039 | 0.603 | 0.405 | 0.805 | 0.503 | 0.025 | 0.574 | 0.645 |
| $\alpha_2$ | 0.359 | 0.323 | 0.241 | 0.618 | 0.637 | 0.453 | 0.662 | 0.901 |
| $\beta$ | 0.214 | 0.295 | 0.722 | 0.7 | 0.684 | 0.268 | 0.599 | 0.562 |
| $\gamma$ | 0.038 | 0.069 | 0.676 | 0.457 | 0.523 | 0.807 | 0.462 | 0.701 |

(b) $\mathcal{X}(\mathcal{RA}) > \mathcal{X}(\mathcal{O})$
| | $BC$ | $c$ | $\mathcal{C}$ | $N$ | $E$ | $\beta_0$ | $\beta_1$ | $\beta_2$ |
| --- | --- | --- | --- | --- | --- | --- | --- | --- |
| $B$ | 0.0 | 0.919 | 0.121 | 0.062 | 0.17 | 0.001 | 0.249 | 0.693 |
| $\delta$ | 0.001 | 0.763 | 0.001 | 0.0 | 0.003 | 0.801 | 0.003 | 0.706 |
| $\theta$ | 0.0 | 0.306 | 0.215 | 0.013 | 0.087 | 0.495 | 0.122 | 0.872 |
| $\alpha_1$ | 0.0 | 0.359 | 0.008 | 0.003 | 0.01 | 0.053 | 0.056 | 0.8 |
| $\alpha_2$ | 0.0 | 0.546 | 0.262 | 0.024 | 0.023 | 0.965 | 0.05 | 0.92 |
| $\beta$ | 0.0 | 0.517 | 0.005 | 0.0 | 0.0 | 0.121 | 0.0 | 0.671 |
| $\gamma$ | 0.0 | 0.331 | 0.0 | 0.0 | 0.0 | 0.525 | 0.0 | 0.958 |

(c) $\mathcal{X}(\mathcal{RA}) > \mathcal{X}(\mathcal{N})$
| | $BC$ | $c$ | $\mathcal{C}$ | $N$ | $E$ | $\beta_0$ | $\beta_1$ | $\beta_2$ |
| --- | --- | --- | --- | --- | --- | --- | --- | --- |
| $B$ | 0.0 | 0.001 | 0.0 | 0.0 | 0.0 | 0.0 | 0.0 | 0.004 |
| $\delta$ | 0.001 | 0.485 | 0.11 | 0.0 | 0.0 | 0.522 | 0.002 | 0.387 |
| $\theta$ | 0.0 | 0.01 | 0.079 | 0.0 | 0.0 | 0.645 | 0.001 | 0.03 |
| $\alpha_1$ | 0.034 | 0.433 | 0.611 | 0.003 | 0.0 | 0.043 | 0.006 | 0.315 |
| $\alpha_2$ | 0.001 | 0.352 | 0.003 | 0.002 | 0.002 | 0.61 | 0.01 | 0.925 |
| $\beta$ | 0.002 | 0.139 | 0.122 | 0.0 | 0.0 | 0.472 | 0.003 | 0.163 |
| $\gamma$ | 0.108 | 0.038 | 0.042 | 0.067 | 0.064 | 0.891 | 0.185 | 0.539 |

TABLE S.5: p-values corresponding to results of figure 4 of the main text.
| | $BC$ | $c$ | $\mathcal{C}$ | $N$ | $E$ | $\beta_0$ | $\beta_1$ | $\beta_2$ |
| --- | --- | --- | --- | --- | --- | --- | --- | --- |
| $B$ | 0.0 | 0.0 | 0.0 | 0.0 | 0.0 | 0.0 | 0.0 | 0.0 |
| $\delta$ | 0.0 | 0.516 | 0.0 | 0.0 | 0.272 | 0.0 | 0.0 | 0.224 |
| $\theta$ | 0.0 | 0.039 | 0.0 | 0.0 | 0.14 | 0.0 | 0.0 | 0.025 |
| $\alpha_1$ | 0.0 | 0.806 | 0.0 | 0.0 | 0.408 | 0.0 | 0.0 | 0.248 |
| $\alpha_2$ | 0.0 | 0.775 | 0.0 | 0.0 | 0.646 | 0.0 | 0.0 | 0.955 |
| $\beta$ | 0.0 | 0.191 | 0.0 | 0.0 | 0.069 | 0.0 | 0.0 | 0.071 |
| $\gamma$ | 0.0 | 0.127 | 0.0 | 0.0 | 0.519 | 0.0 | 0.0 | 0.531 |

### S.6. MULTI-FREQUENCY ANALYSIS: INDIVIDUAL PATIENT RESULTS

In this supplementary section we detail the definition of the distinguishability score *D* and provide details on the statistical analyses involving this metric. In Supp. Figure S.3 we show the results of the node-based analyses. Each panel corresponds to a frequency band and a comparison between node-sets, as indicated by the panel title. For each panel, we show the result 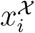 of the statistical comparison between the two node-sets, using each of the centrality metrics 𝒳 and for each patient *i* with a color code. The color code indicates whether there is a significant difference in the direction of the hypothesis (blue, 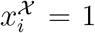), against it (red, 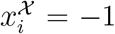), or there is no significant difference (grey, 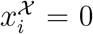). The patient distinguishability score *D*_*i*_ is simply defined as the sum of the results of this statistical comparison over generalized centrality metrics: 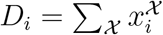. Given that central nodes have large negative curvature, this term is multiplied by −1 in the sum. The resulting patient distinguishability score *D*_*i*_ is thus a number between −8 and 8, where *D*_*i*_ = 8 (−8) indicates that the two node-sets were highly different in the direction of the hypothesis (against the hypothesis), and *D*_*i*_ = 0 indicates no significant or inconsistent differences (across metrics) for the patient.

In Supp. Figure S.4 we show the results of the statistical comparison between the SF and NSF groups based on the distinguishability scores *D*_*i*_, for each of the node-based tests. We observed a tendency towards higher scores for SF patients for the broadband, *θ* and *α*_1_ bands, and in the opposite direction for *δ* and *γ*, however none of the differences are significant after Bonferroni correction for multiple comparisons.

Finally, we also performed a receiver-operating-characteristic (ROC) patient-classification analysis based on the distinguishability scores, the results of which were reported in the main text. Here we show in Supp. Figure S.5 the ROC curves corresponding to each of the node-based tests, for the broadband.

**FIG. S.3:**
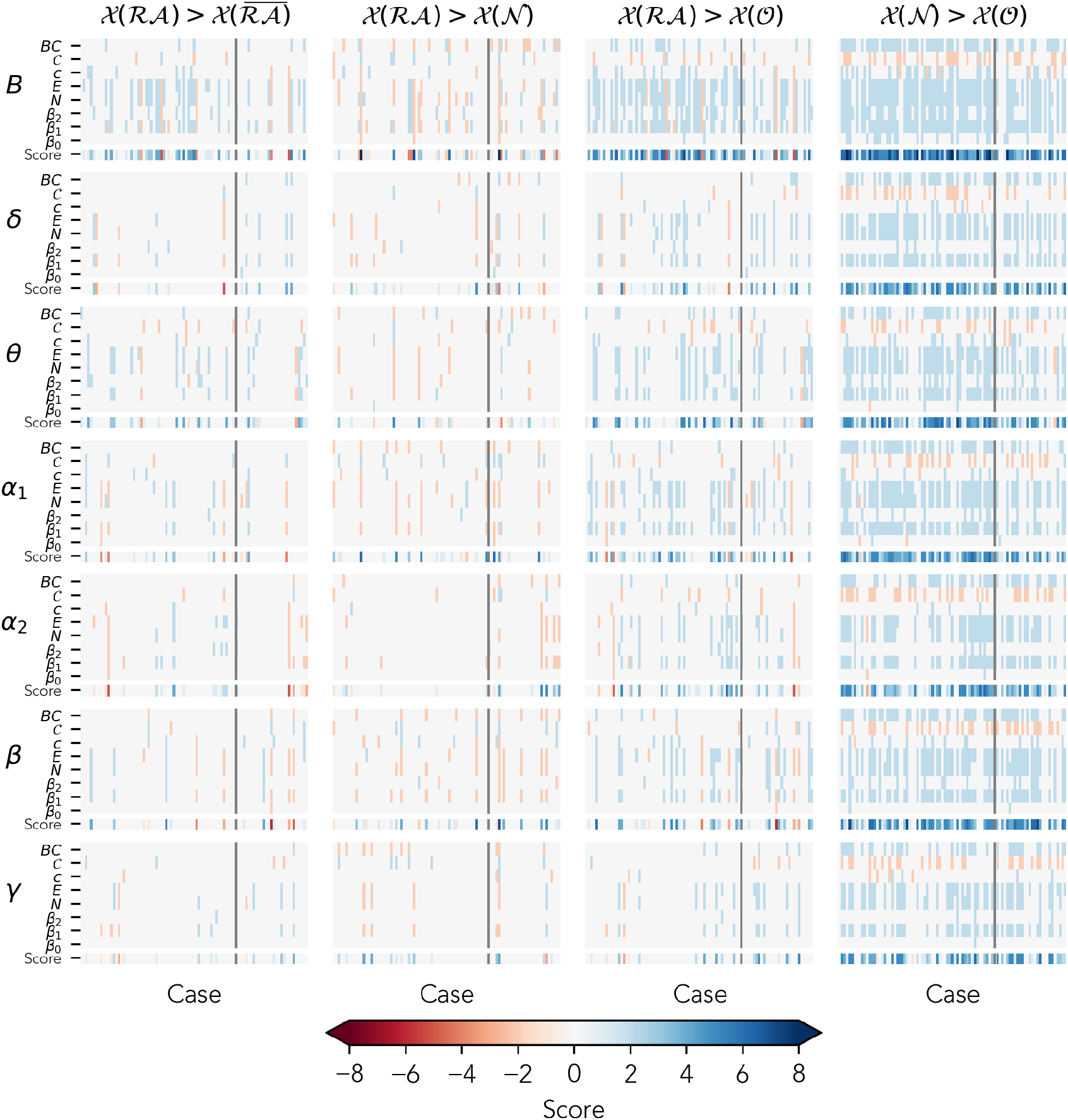
Disthinguishability score *D*_*i*_. Each panel corresponds to the comparison between two node-sets as indicated by the panel titles, and a frequency band (from top to bottom: broadband, *δ, θ, α*_1_, *α*_2_, *β, γ*). We show the result for the statistical comparison 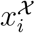 for each metric 𝒳 and each patient *i* in the top rows of each panel and *D*_*i*_ in the bottom row. All metrics are color-coded as indicated by the colorbar. The vertical grey line on each panel separate SF (left) and NSF (right) cases.

**FIG. S.4:**
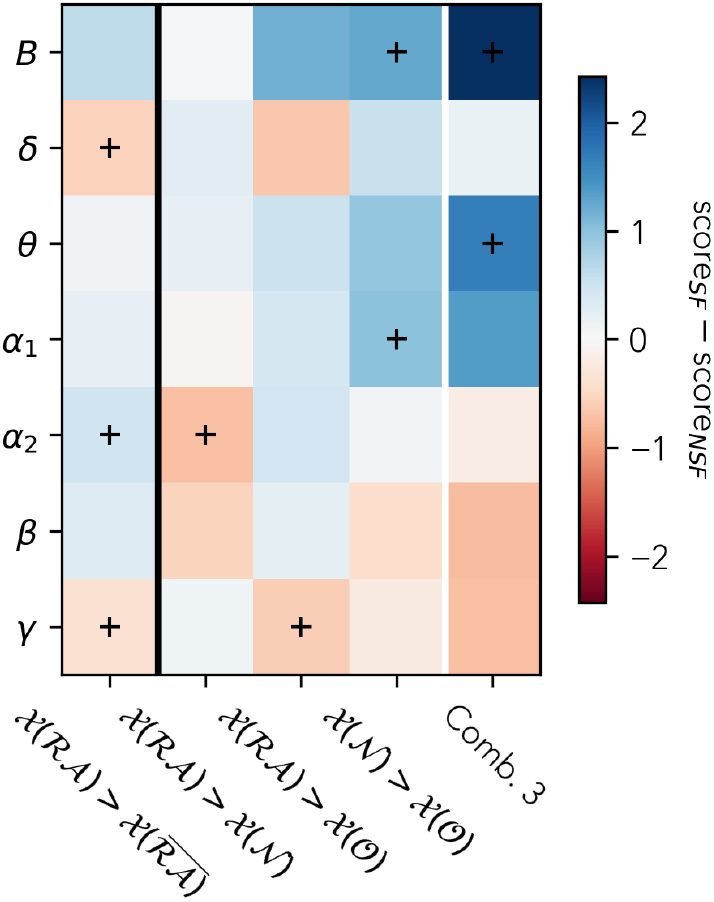
Statistical comparison between the SF and NSF patient groups based on the patient distinguishability score *D*_*i*_, for each node-group-comparison (columns) and frequency band (rows), as indicated by the axis labels. The final column combines the results of the three-node-groups tests by adding up the patient scores. The color-code indicates the difference between the average scores of the SF and NSF groups, as given by the color-bar. The cross markers indicate differences with p-value *<* 0.05 before Bonferroni correction for multiple comparisons. None of the differences were significant after the correction.

### S.7. ALTERNATIVE DISTINGUISHABILITY SCORE

In order to compare our findings with a recent study by Ramaraju and colleagues [43], we repeated the patient-classification analysis using their original definition of the distinguishability score, 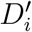. We also consider the same centrality metric used by [43], the weighted degree or strength of a node (the sum of its link weights after thresholding the PLI matrix with the disparity filter). The distinguishability score for each patient 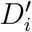 was defined by [43] as the AUC of the ROC-classification analysis of the ℛ𝒜 and 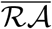 node sets. A *D*^*′*^ value close to 0.5 indicates that the two node sets cannot be classified according to the corresponding metric, whereas values close to 0 or 1 indicate that the node sets are easily classifiable. In particular, *AUC >* 0.5 indicates that the ℛ𝒜 set is more central than the 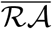 set, and vice versa for *AUC <* 0.5. The results of this analysis are shown in Supp. Table S.6 (first column).

**FIG. S.5:**
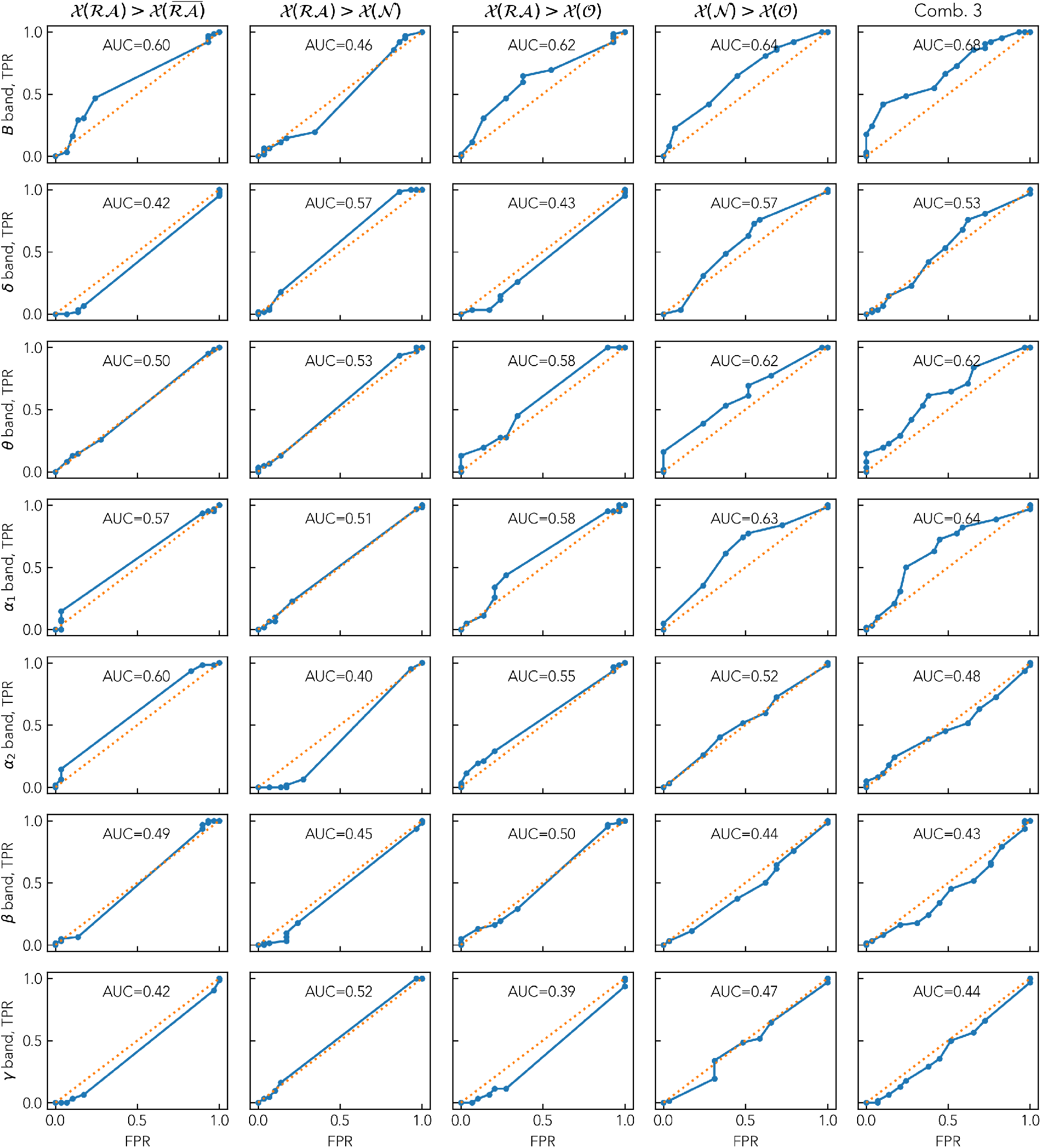
ROC analysis of the SF and NSF groups based on the patient scores, for each frequency band (rows) and node-groups analysis (columns). The final column corresponds to the compounded score of the three-node-group analysis. We indicate the area under the curve (AUC) of each curve as the legend.

We did not find a good patient classification for any frequency band. The best classification results were obtained for the *γ* band with *AUC* = 0.64, followed by the *α*_1_ (*AUC* = 0.39) and *α*_2_ (*AUC* = 0.60) bands. Interestingly, the direction of the classification changed across frequency bands: for *α*_2_ and *γ* SF patients presented higher distinguishability scores 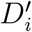 than NSF patients, whereas for *α*_1_ the opposite was true.

In order to exploit the three-node-set partition framework defined in the main text, we extended this analysis to account for three more two-class node-based classifications, namely i) ℛ𝒜 and 𝒩 nodes; ii) ℛ𝒜 and 𝒪 nodes; and 𝒩 and 𝒪 nodes (Supp. Table S.6). Swarm plots depicting distinguishability scores are presented in Supp. figure S.6 for the case of broad band for a visual representation of the classification. We found that the results for the latter two cases were very similar to the original ℛ𝒜 and 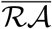 distinguishability. As expected from the results in Figure 3 in the main text, the ℛ𝒜 and 𝒩 cannot be easily classified, resulting in low node-distinguishability scores and in a poor patient classification.

**TABLE S.6:** Results of the patient classification following the methodology in [43]. We report the area under the curve (AUC) of the patient classification (SF versus NSF) based on the distinguishability *D*^*′*^ between ℛ𝒜 and 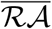 nodes (first column), when using the weighted degree as nodal centrality metric. 𝒳 (*S*) stands for the generalized centrality metric 𝒳 measured on the nodes in set *S*. The latter three columns extend this analysis to the three-node-set framework by considering the distinguishability between the i) ℛ𝒜 and *N* nodes; ii) ℛ𝒜 and 𝒪 nodes; and 𝒩 and 𝒪 node sets, respectively. Each row corresponds to a different frequency band. We highlight in bold the results for with |*AUC* − 0.5| *>* 0.1.

| | $\mathcal{X}(\mathcal{RA}) > \mathcal{X}(\overline{\mathcal{RA}})$ | $\mathcal{X}(\mathcal{RA}) > \mathcal{X}(\mathcal{N})$ | $\mathcal{X}(\mathcal{RA}) > \mathcal{X}(\mathcal{O})$ | $\mathcal{X}(\mathcal{N}) > \mathcal{X}(\mathcal{O})$ |
| --- | --- | --- | --- | --- |
| $B$ | 0.56 | 0.51 | 0.57 | 0.49 |
| $\delta$ | 0.57 | 0.56 | 0.59 | 0.57 |
| $\theta$ | 0.42 | 0.46 | 0.43 | 0.44 |
| $\alpha_1$ | **0.39** | 0.41 | **0.40** | 0.51 |
| $\alpha_2$ | **0.60** | 0.55 | **0.61** | **0.60** |
| $\beta$ | 0.48 | 0.47 | 0.45 | 0.50 |
| $\gamma$ | **0.64** | 0.58 | **0.65** | 0.57 |

**FIG. S.6:**
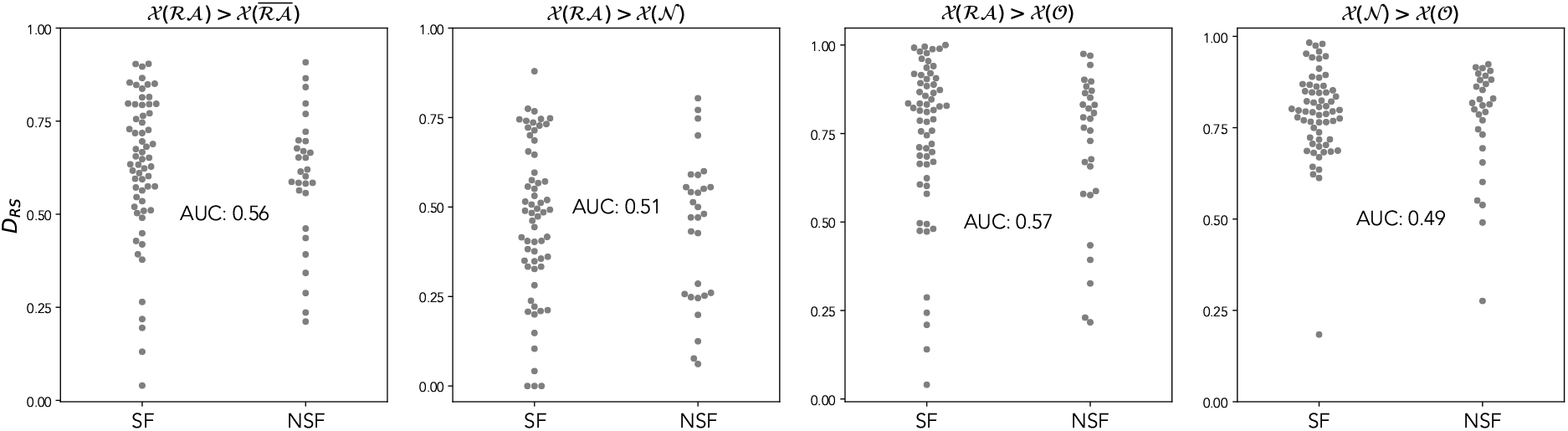
Swarm plot depicting distinguishability values 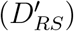 for SF and NSF surgical outcomes for different node-based comparisons, based on the node strength. 𝒳 (*S*) stands for the generalized centrality metric 𝒳 (node strength here) measured on the nodes in set *S*. Values close to 0 (1) indicate that high strength nodes are resected (spared). Each scatter point represents an individual patient. The results for every band are reported in Table S.6.

Finally, we repeated this analysis on our proposed framework of 8 generalized centrality metrics, the results are shown in Supp. Figure S.7. The results were similar to those using the weighted degree, with only fair patient classification results. The best findings were obtained when considering the two-node-set partition (i.e. ℛ𝒜 versus 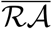) in the broadband (*AUC* = 0.68 for the metric *E*), and overall showed large variability also in the direction of the *AUC* (that is, whether SF or NSF patients presented larger distinguishability scores). Thus this extended analysis was not able to improve upon our initial results.

**FIG. S.7:**
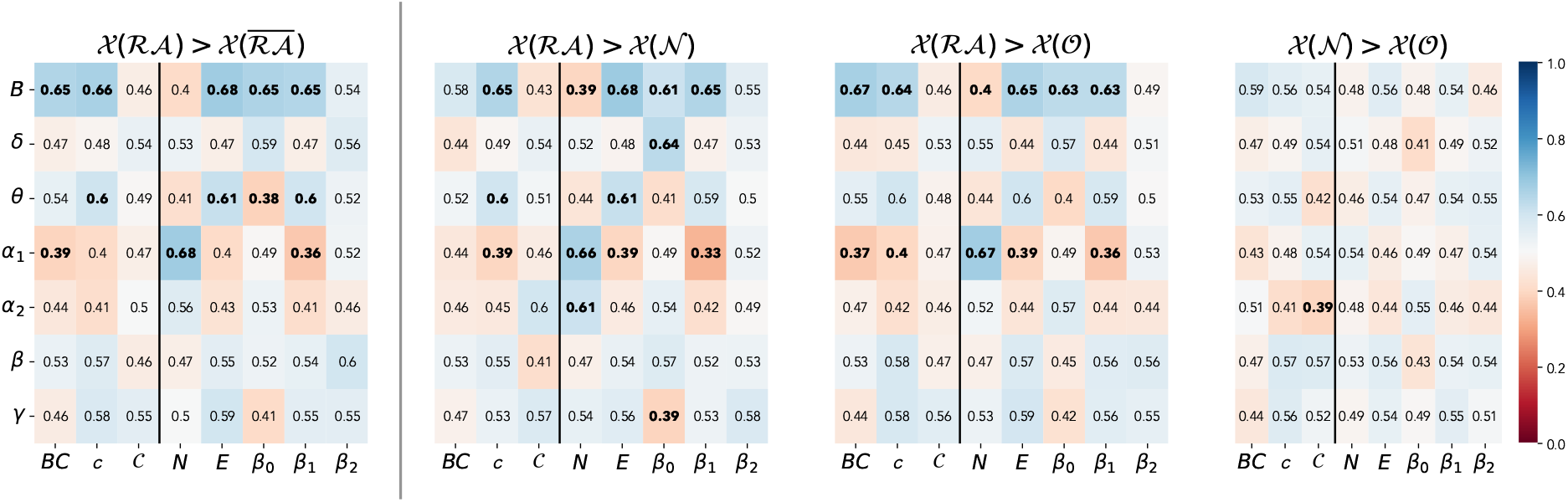
Patient classification results using the methodology by [43] combined with our proposed generalized centrality metrics. Each panel corresponds to a node-based comparison as indicated by the panel titles, with the vertical line separating the two-node-set case from the three-node-set cases. 𝒳(*S*) stands for the generalized centrality metric 𝒳 measured on the nodes in set *S*. Rows correspond to frequency bands and columns to generalized centrality metrics. We show the resulting AUC both with the color-code and by numerical values. Bold numbers correspond to |*AUC* − 0.5| *>* 0.1.

